# Estimating the Population Benefits of Blood Pressure Lowering: A Wide-Angled Mendelian Randomization Study in UK Biobank

**DOI:** 10.1101/2021.02.02.21250515

**Authors:** Hannah Higgins, Amy M. Mason, Susanna C. Larsson, Dipender Gill, Claudia Langenberg, Stephen Burgess

**Affiliations:** Department of Public Health and Primary Care, University of Cambridge, Cambridge, UK; Department of Surgical Sciences, Uppsala University, Uppsala, Sweden; Unit of Cardiovascular and Nutritional Epidemiology, Institute of Environmental Medicine, Karolinska Institutet, Stockholm, Sweden; Department of Epidemiology and Biostatistics, School of Public Health, Imperial College London, London, UK; Centre for Pharmacology and Therapeutics, Department of Medicine, Hammersmith Campus, Imperial College London, London, UK; Department of Genetics, Novo Nordisk Research Centre Oxford, Old Road Campus, Oxford, UK; Clinical Pharmacology and Therapeutics Section, Institute of Medical and Biomedical Education and Institute for Infection and Immunity, St George’s, University of London, London, UK; Clinical Pharmacology Group, Pharmacy and Medicines Directorate, St George’s University Hospitals NHS Foundation Trust, London, UK; MRC Epidemiology Unit, University of Cambridge, Cambridge, UK; Medical Research Council Biostatistics Unit, University of Cambridge, Cambridge, UK

**Keywords:** Cardiovascular disease, Mendelian randomization, high blood pressure, hypertension, genetic epidemiology

## Abstract

**Background:** The causal relevance of elevated blood pressure for several cardiovascular diseases is uncertain, as is the population impact of blood pressure lowering on risk of cardiovascular diseases more broadly. This study systematically assesses evidence of causality for various cardiovascular diseases in a two-sample Mendelian randomization framework, and estimates the potential reduction in the prevalence of these diseases attributable to long-term population shifts in the distribution of systolic blood pressure (SBP).

**Methods and Results:** We investigated associations of genetically-predicted SBP as predicted by 256 genetic variants with 21 cardiovascular diseases in UK Biobank, a population-based cohort of UK residents. The sample consisted of 376,703 participants of European ancestry aged 40-69 years at baseline. Genetically-predicted SBP was positively associated with 14 of the outcomes (p<0.002), including dilated cardiomyopathy, endocarditis, peripheral vascular disease, and rheumatic heart disease. Using genetic variation to estimate the long-term impact of blood pressure lowering on disease, population reductions in SBP were predicted to result in an overall 16.9% (95% confidence interval (CI): 12.2-21.3%) decrease in morbidity for a 5 mmHg decrease from a population mean of 137.7 mmHg, 30.8% (95% CI: 22.8-38.0%) for a 10 mmHg decrease, and 56.2% (95% CI: 43.7-65.9%) decrease for a 22.7 mmHg decrease in SBP (22.7 mmHg represents a shift from the current mean SBP to 115 mmHg).

**Conclusions:** Risk of many cardiovascular diseases is influenced by long-term differences in SBP. The burden of a broad range of CVDs could be substantially reduced by long-term population-wide reductions in the distribution of blood pressure.

## Introduction

High blood pressure has severe, costly consequences largely through increased cardiovascular disease (CVD) risk ^1^. For many CVDs, a causal relationship with blood pressure that is reversible through treatment has been demonstrated in randomized controlled trials (RCTs) ^2^. However, for diseases such as dilated cardiomyopathy, endocarditis, peripheral vascular disease, and aortic valve stenosis, RCT evidence demonstrating a causal effect of blood pressure lowering is lacking. Despite this, these outcomes have been used to estimate the population impact of raised blood pressure ^3^. Additionally, quantitative evidence for the benefit of reducing blood pressure has not been assessed for several CVD outcomes in a primary prevention setting.

In the absence of RCT evidence, Mendelian randomization (MR) can circumvent several limitations of observational epidemiology, allowing unconfounded inferences from observational data ^4^. MR uses selected genetic variants related to an exposure to provide evidence supporting a causal hypothesis. The independent segregation of alleles at conception means that genetically-defined subgroups of the population with increased or decreased average blood pressure levels should not differ systematically with respect to confounding variables, creating a natural experiment analogous to an RCT ^4^. Life-long average differences in the exposure between subgroups compared in an MR analysis provide evidence regarding the potential impact of long-term interventions on the exposure, in contrast to the short-term interventions usually evaluated by RCTs ^5^.

Here, we employ MR to assess evidence for causality between systolic blood pressure (SBP) and a broad range of CVDs in UK Biobank, a population-based cohort of UK residents. We then use these estimates to predict the potential reduction in cardiovascular disease burden in the UK population attributable to distributional shifts in SBP. We focus on SBP as a measure of blood pressure because it is a better predictor of health outcomes than diastolic blood pressure (DBP) ^1^. However, as the genetic variants used in this investigation are associated with both SBP and DBP, estimates relate generally to blood pressure lowering and are not specific to SBP.

## Methods

We performed two-sample Mendelian randomization analyses using summarized data (eFigure 1). Genetic associations with blood pressure were obtained in 299,024 European ancestry participants from the International Consortium for Blood Pressure (ICBP) ^6^. Genetic associations with 21 CVDs were estimated in 376,703 European ancestry participants from UK Biobank ^7^ by logistic regression with adjustment for age, sex, and 10 genomic principal components (eTable 1). As the ICBP and UK Biobank samples do not overlap, bias due to winner’s curse is avoided. As genetic instruments, we selected 256 variants previously associated with blood pressure at a genome-wide level of significance in the ICBP dataset (eTable 2). The variants explained 2.1% of variance in SBP in ICBP, corresponding to an F-statistic of 23.1. Associations between a weighted genetic risk score (GRS) and potential confounders (sex, age, BMI, smoking status, low-density lipoprotein [LDL] cholesterol, alcohol drinker status, and anti-hypertensive medication status) were assessed in UK Biobank.

**Table 1.**
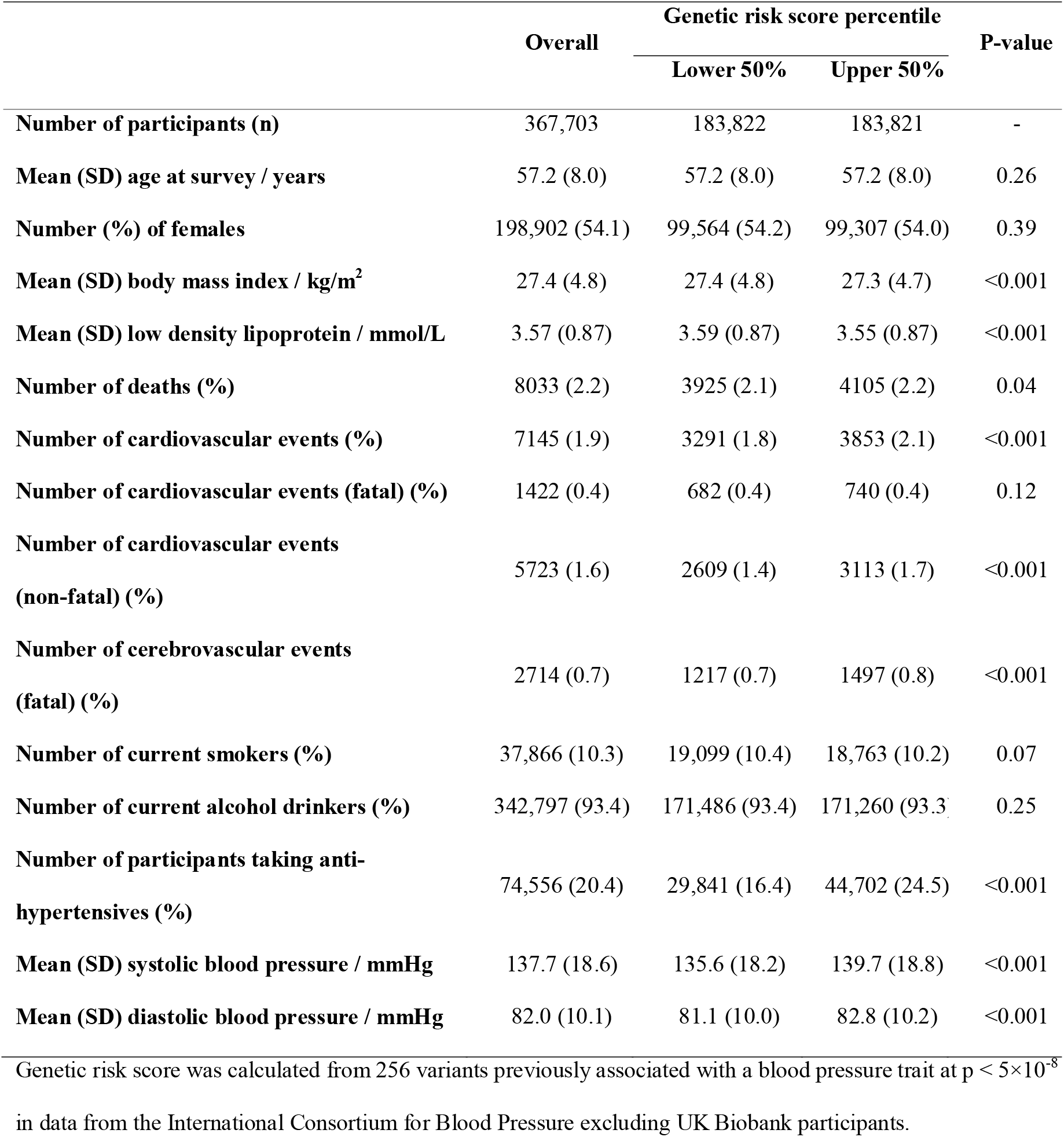
Baseline characteristics of participants in the analytic subset of the UK Biobank study

|  | Overall | Genetic risk score percentile |  | P-value |
| --- | --- | --- | --- | --- |
|  |  | Lower 50% | Upper 50% |  |
| **Number of participants (n)** | 367,703 | 183,822 | 183,821 | - |
| **Mean (SD) age at survey / years** | 57.2 (8.0) | 57.2 (8.0) | 57.2 (8.0) | 0.26 |
| **Number (%) of females** | 198,902 (54.1) | 99,564 (54.2) | 99,307 (54.0) | 0.39 |
| **Mean (SD) body mass index / kg/m<sup>2</sup>** | 27.4 (4.8) | 27.4 (4.8) | 27.3 (4.7) | <0.001 |
| **Mean (SD) low density lipoprotein / mmol/L** | 3.57 (0.87) | 3.59 (0.87) | 3.55 (0.87) | <0.001 |
| **Number of deaths (%)** | 8033 (2.2) | 3925 (2.1) | 4105 (2.2) | 0.04 |
| **Number of cardiovascular events (%)** | 7145 (1.9) | 3291 (1.8) | 3853 (2.1) | <0.001 |
| **Number of cardiovascular events (fatal) (%)** | 1422 (0.4) | 682 (0.4) | 740 (0.4) | 0.12 |
| **Number of cardiovascular events (non-fatal) (%)** | 5723 (1.6) | 2609 (1.4) | 3113 (1.7) | <0.001 |
| **Number of cerebrovascular events (fatal) (%)** | 2714 (0.7) | 1217 (0.7) | 1497 (0.8) | <0.001 |
| **Number of current smokers (%)** | 37,866 (10.3) | 19,099 (10.4) | 18,763 (10.2) | 0.07 |
| **Number of current alcohol drinkers (%)** | 342,797 (93.4) | 171,486 (93.4) | 171,260 (93.3) | 0.25 |
| **Number of participants taking anti-hypertensives (%)** | 74,556 (20.4) | 29,841 (16.4) | 44,702 (24.5) | <0.001 |
| **Mean (SD) systolic blood pressure / mmHg** | 137.7 (18.6) | 135.6 (18.2) | 139.7 (18.8) | <0.001 |
| **Mean (SD) diastolic blood pressure / mmHg** | 82.0 (10.1) | 81.1 (10.0) | 82.8 (10.2) | <0.001 |

3 Genetic risk score was calculated from 256 variants previously associated with a blood pressure trait at $p < 5 \times 10^{-8}$ 4 in data from the International Consortium for Blood Pressure excluding UK Biobank participants.

**Figure 1.**
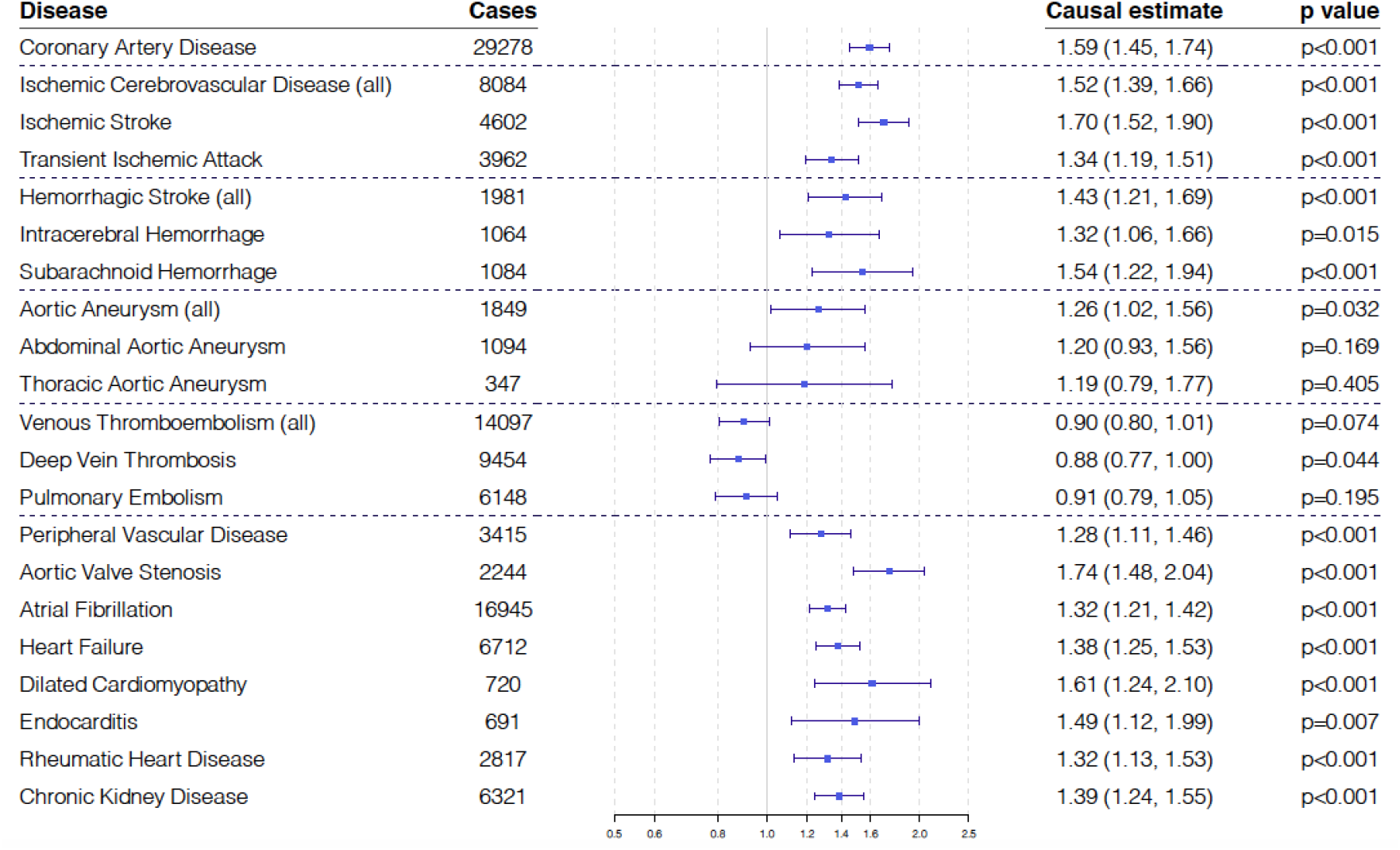
Mendelian randomization estimates (odds ratio with 95% confidence interval per 10 mmHg increase in genetically-predicted SBP) using the inverse-variance weighted method.

Mendelian randomization estimates for the casual effect of SBP on the odds of 21 CVD outcomes were obtained using the inverse-variance weighted (IVW) method ^8^. Estimates are odds ratios per 10 mmHg increase in genetically-predicted SBP. Sensitivity analyses were performed using the MR-Egger, weighted median, and MR-PRESSO methods ^8^. Outcomes with strong evidence for causality (p-value < 0.05/21 = 0.002 in either IVW or MR-PRESSO method and concordance of estimates across all methods) were used to estimate the change in disease burden that would occur under interventions in the distribution of SBP. Assuming a linear model, we estimated the population impact fraction ^3^, representing the relative reduction in disease risk if mean SBP were set to a given value for all individuals in the population. We also estimated the absolute reduction in events from a population shift in the distribution of SBP using disease prevalence estimates from surveys relevant to a middle-to early late-aged UK-based population (eTable 3).

## Results

SBP was approximately normally distributed with a mean of 137.7 mmHg (standard deviation 18.6 mmHg) (eFigure 2). There was no association between the GRS and age, sex, smoking, or alcohol consumption (Table 1). The GRS was associated with BMI and with LDL cholesterol (mean difference between top versus bottom 50%: 4.1 mmHg for SBP, -0.1 kg/m^2^ for BMI, -0.04 mmol/L for LDL-cholesterol). Hence any influence of BMI or LDL-cholesterol on estimates via pleiotropy should be minimal, and would generally result in underestimation of the effect of SBP as both BMI and LDL-cholesterol increase the risk of most CVDs.

**Figure 2.**
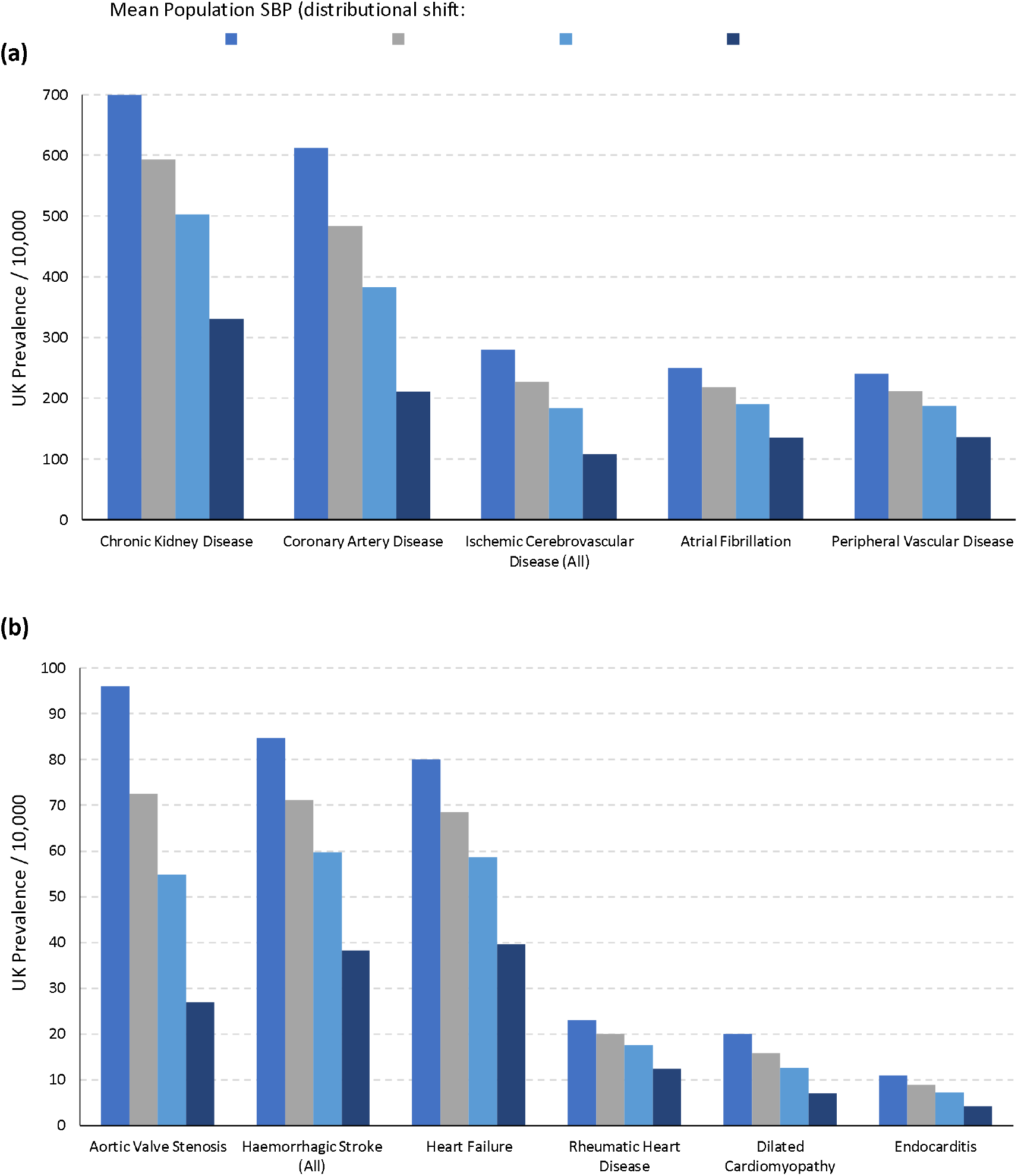
(a-b) Bar chart showing the prevalence and estimated changed if SBP decreased across its distribution by 5 mmHg, 10 mmHg, and 22.7 mmHg (22.7 mmHg represents a shift from the current mean of 137.7 mmHg to 115 mmHg) separately for each cardiovascular outcome and for **(a)** high prevalence outcomes (current UK prevalence > 200 per 10,000) and **(b)** lower prevalence outcomes (current UK prevalence < 200 per 10,000). Ischemic cerebrovascular disease comprises ischemic stroke and transient ischemic attack. Hemorrhagic stroke comprises subarachnoid hemorrhage and intracerebral hemorrhage.

Fourteen outcomes satisfied the criteria of strong evidence for causality: p< 0.002 and concordance of estimates across methods (Figure 1, eTable 4). In decreasing order of the IVW estimate, these were: aortic valve stenosis, ischemic stroke, dilated cardiomyopathy, coronary artery disease, subarachnoid hemorrhage, ischemic cerebrovascular disease, endocarditis, hemorrhagic stroke (all), chronic kidney disease, heart failure, transient ischemic attack, atrial fibrillation, rheumatic heart disease, and peripheral vascular disease (eFigure 3). Two further outcomes (intracerebral hemorrhage and aortic aneurysm) had a positive IVW estimate at a conventional level of statistical significance (p<0.05). Deep vein thrombosis had an inverse IVW estimate at a conventional level of statistical significance (p<0.05).

Figure 2 shows the estimated changes in the absolute prevalence of outcomes with strong evidence for causality resulting from a population shift in the distribution of SBP. Population impact fractions for these outcomes are provided in eTable 5. Aggregating across these outcomes, reductions in SBP were predicted to result in an overall 16.9% (95% confidence interval (CI): 12.2-21.3%) decrease in CVD morbidity for a 5 mmHg SBP decrease from a population mean of 137.7 mmHg, 30.8% (95% CI: 22.8-38.0%) for a 10 mmHg SBP decrease, and 56.2% (95% CI: 43.7-65.9%) decrease for a 22.7 mmHg SBP decrease 22.7 mmHg represents a shift from the current mean SBP in the population to 115 mmHg, a value that has been proposed as a theoretical minimum risk target.

## Discussion

While for many CVDs the causal effect of blood pressure lowering has been demonstrated convincingly in RCTs, several outcomes (dilated cardiomyopathy, endocarditis, peripheral vascular disease, and rheumatic heart disease) had previously only been shown to be associated with SBP in observational studies ^1,3^. Our analysis adds evidential weight to blood pressure as a causal risk factor for these outcomes and to their inclusion in other SBP population impact studies.

Although the majority of the outcomes associated with genetically-predicted SBP are chronic diseases, some have infectious origins (for example, endocarditis and rheumatic heart disease). Elevated SBP may therefore increase susceptibility to, or damage from, infection. A notable finding was the inverse association between genetically-predicted SBP and deep vein thrombosis. Although this association did not achieve statistical significance after accounting for multiple testing, an inverse association between SBP and venous thromboembolism has been found in previous observational studies ^9,10^.

The MR estimates obtained here were generally larger than those from RCTs. For example, the MR estimate for the risk of ischemic stroke per 10 mmHg increase in SBP was 1.70 (95% CI: 1.52-1.90). This contrasts with an estimate of 1.27 (95% CI: 1.23-1.32) from a recent meta-analysis of RCTs ^2^. While this is expected as MR estimates represent the lifelong impact of elevated blood pressure whereas RCTs vary blood pressure for a shorter period ^11^, other factors such as trial setting and inclusion criteria may also contribute.

This study has many strengths, but also limitations. The large sample size and wide range of outcomes enables systematic cross-comparisons of unconfounded estimates in a single cohort. This is important from a public health perspective when comparing the impact of the same genetic change on different diseases. The genome-wide genotypic data available from the cohort absolved any need to use proxy SNPs in the instrument. However, the results should be interpreted in the context of several limitations.

First, the UK Biobank cohort is somewhat unrepresentative of the UK population and suffers from ‘healthy volunteer’ selection bias ^12^. Analyses were conducted in participants of European descent to avoid population stratification. A recent GWAS identified only modest correlation of blood pressure effect estimates across ancestries ^13^. Estimates may therefore not be fully relevant for the whole UK population. Whether the findings are applicable to other ethnicities warrants investigation, particularly since hypertension disproportionately affects black African and Caribbean ancestry ethnic groups both in the UK and abroad ^14^. Given large global disparities in hypertension prevalence (both by country and ancestry ^15^), generalizability beyond the UK of the public health modelling may be limited.

The MR approach is underpinned by assumptions that cannot be completely empirically validated ^4^. However, the GRS for SBP was not strongly associated with major confounders and estimates were generally similar across robust methods. Furthermore, the relevance assumption was fulfilled: all SNPs were significantly associated with SBP and the entire genetic instrument explained 2.1% of variance in SBP in the ICBP GWAS, corresponding to an F-statistic of 23.1.

Although the principles of MR seek to emulate an RCT, the approach differs fundamentally in certain aspects which are relevant when using MR in public health modelling. Firstly, the results presented here reflect lifelong differences in SBP relating to genetic variants that are determined at conception. The reversibility of these long-term effects is unknown, however, reversibility is assumed in the population impact fraction calculations. Whilst the SBP-associated risks of most cardiovascular outcomes have demonstrated reversibility from RCTs, it is unknown whether this applies to the full spectrum of outcomes studied here. There may be no existing intervention applicable to a mature cohort which can imitate the genetic effect, and if such an intervention does exist, the time-lag or age target required to produce the predicted effects is unknown ^4^.

Finally, these analyses assume linearity of effects ^1^. Estimates are likely to be reliable for shifts in SBP of similar magnitude to the genetic associations with SBP, which are around 8-10 mmHg. However, the appropriateness of extrapolation to larger changes in SBP cannot be assessed in this current study.

The evidence presented here suggests that incidence of diseases later in life is influenced by long-term, lifelong differences in SBP exposure, even in lower-risk populations. Therefore, confining interventions to older age groups and individuals over a certain risk threshold will only partially address the totality of disease burden. Interventions that may replicate the lifelong reduction in exposure to elevated SBP evaluated here include non-pharmalogic interventions, such as physical activity, weight control, and sodium reductions, and community-wide programmes, such as consumer awareness campaigns and industry collaboration for food reformulation.

In conclusion, reducing systolic blood pressure by 10 mmHg could reduce the overall burden of a broad range of CVDs by around 30%. These findings contribute to an ever-expanding body of evidence advocating targeted and population-based strategies for management of high blood pressure across the life course.

## Data Availability

Individual-level data are available on application to any bona fide researcher.

## Acknowledgements

This research was conducted using the UK Biobank study under Application Number 29202.

