## Supplementary Tables and Figures for "Estimating the Population Benefits of Blood Pressure Lowering: A Wide-Angled Mendelian Randomization Study in UK Biobank"

**Online Data Supplement**

eFigure 1……………………………………………………………….……………...........2
eFigure 2……………………………………………………………….……………...........3
eFigure 3……………………………………………………………….……………...........4
eTable 1……………………………………………………………….……………............7
eTable 2……………………………………………………………….……………............8
eTable 3……………………………………………………………….……………...........14
eTable 4……………………………………………………………….……………...........15
eTable 5……………………………………………………………….……………...........16
Supplemental References……………………………………………….……………..........17

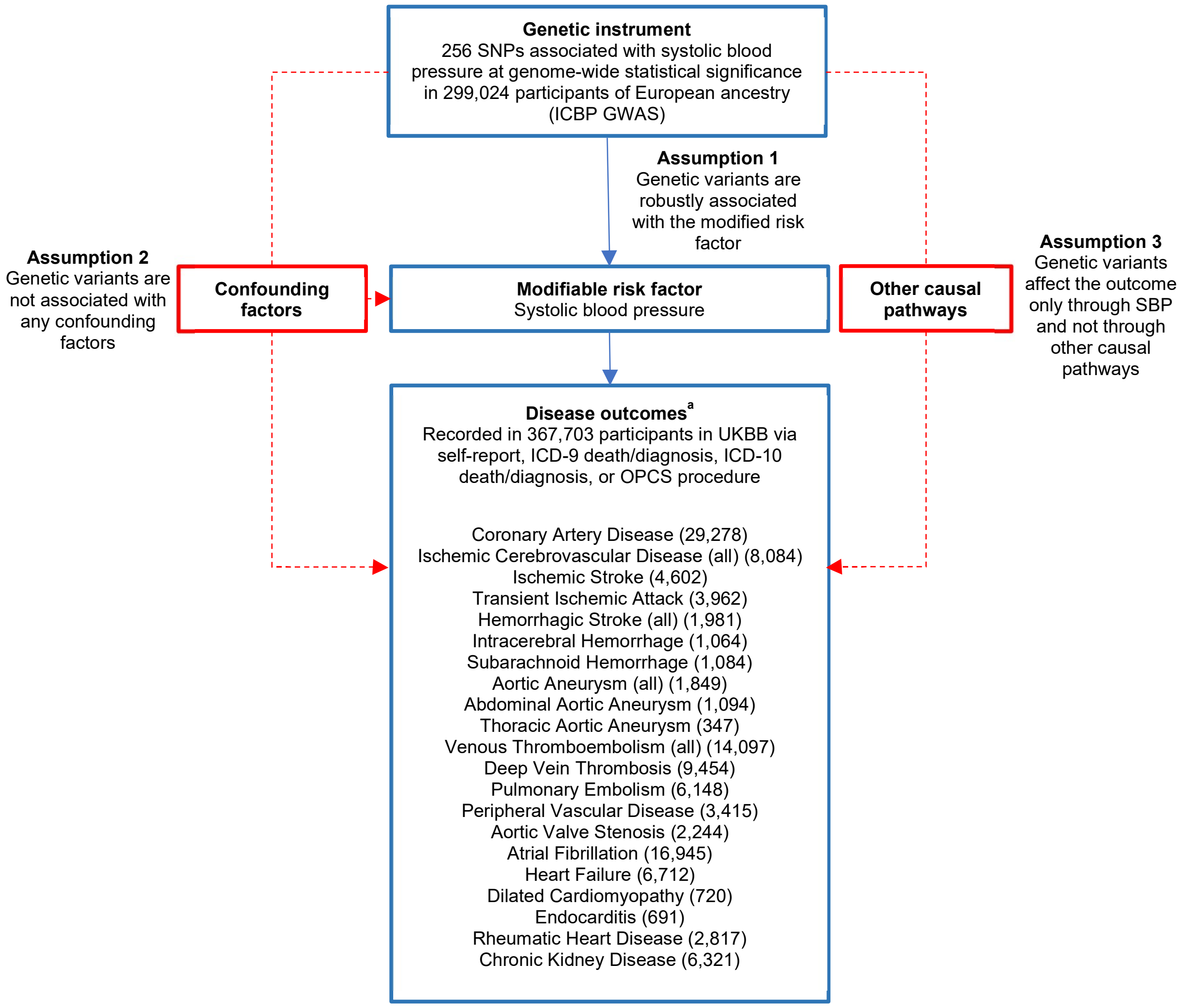


**eFigure 1. Summary of the data sources for this study and the assumptions of the Mendelian randomization design. Broken lines represent potential pleiotropic or direct causal effects between variables that would violate the Mendelian randomization assumptions. GWAS, Genome-wide association study; International Blood Pressure Consortium. ICBP; UKBB, UK Biobank. [a] The number of cases for each outcome is reported in parentheses.**

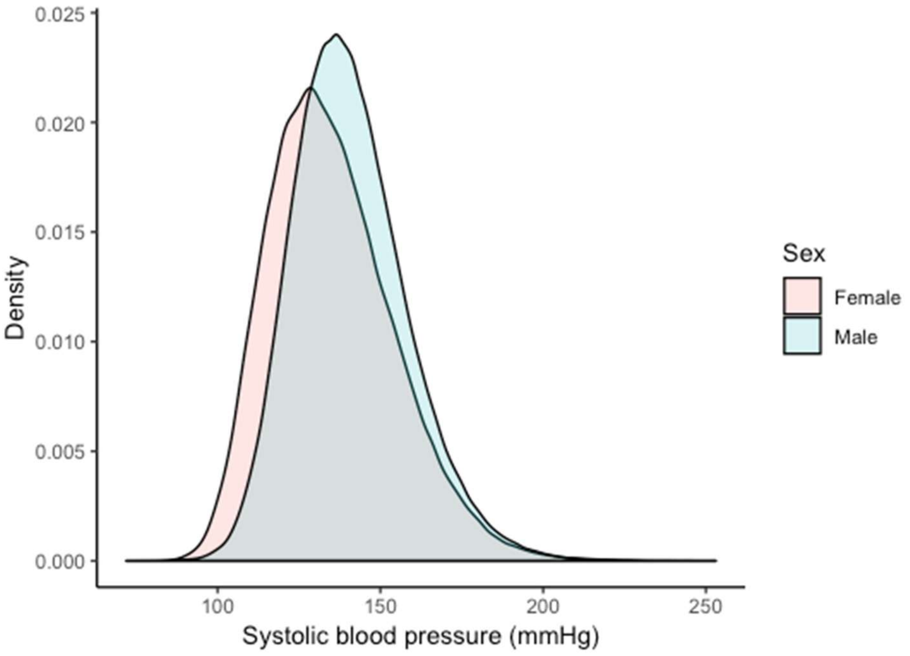


**eFigure 2. Distribution of systolic blood pressure in UK Biobank. Overall mean SBP is 137.7 mmHg (standard deviation 18.6 mmHg); female mean SBP is 135.0 mmHg (standard deviation 19.2); male mean SBP is 140.8 (standard deviation is 17.4 mmHg).**

**(a)**

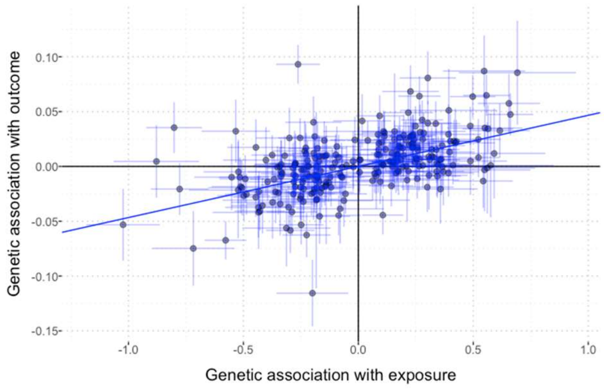


**(b)**

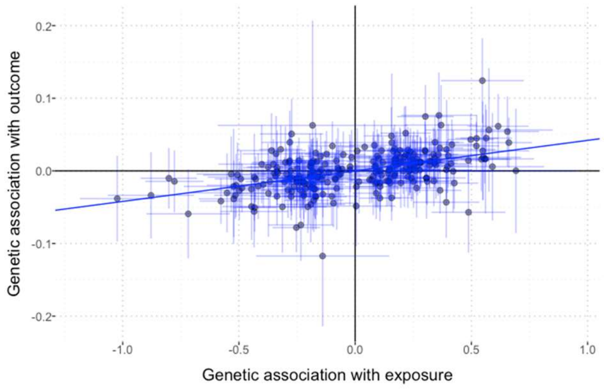


**(c)**

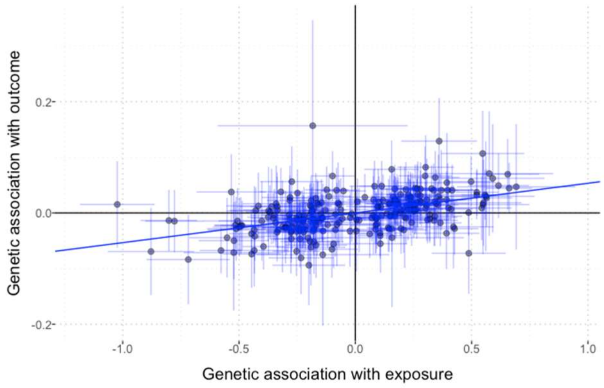


**(d)**

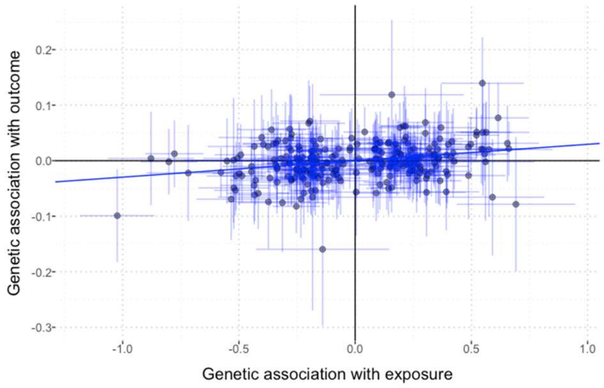


**(e)**

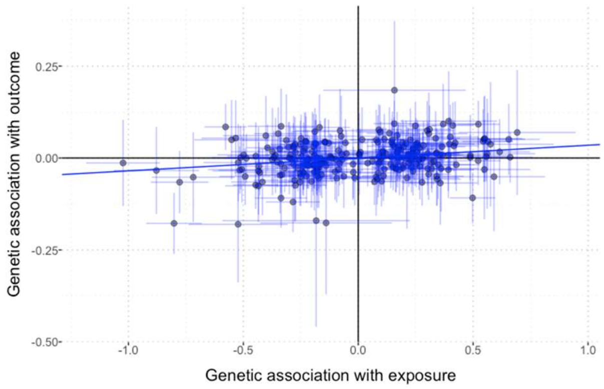


**(f)**

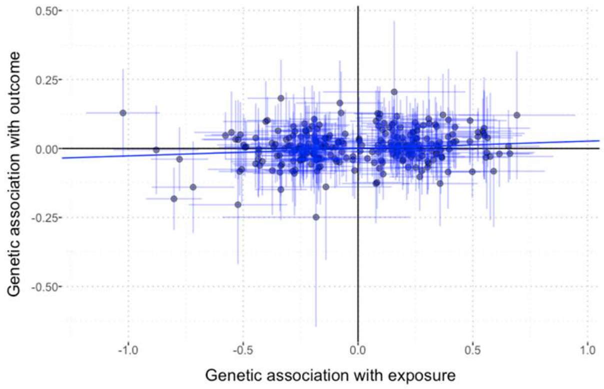


**(g)**

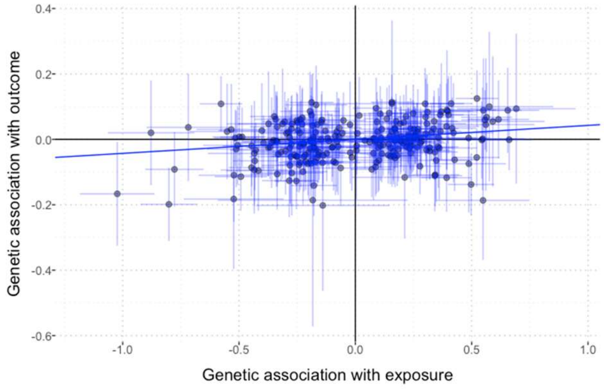


**(h)**

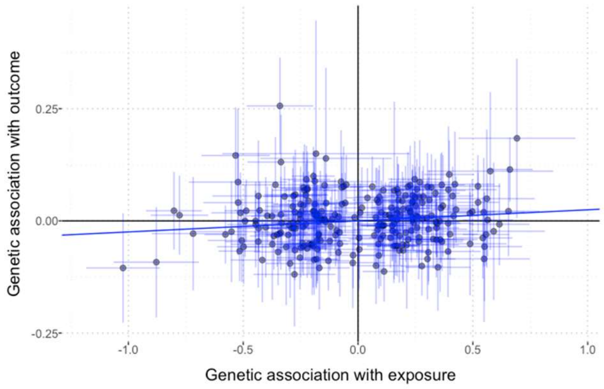


**eFigure 3 (a)-(m). Scatter plots of the beta-coefficients for the exposure plotted against the beta-coefficients for the outcome for each of the 256 SNPs in the instrument. Each dot corresponds to a SNP. Lines represent the 95% CI for each SNP's beta-coefficient. Each scatter plot corresponds to one outcome: (a) Coronary artery disease; (b) Ischemic cerebrovascular disease (all); (c) Ischemic stroke; (d) Transient ischemic attack; (e) Hemorrhagic stroke (all); (f) Intracerebral hemorrhage; (g) Subarachnoid hemorrhage; (h) Aortic aneurysm (all); (i) Abdominal aortic aneurysm; (j) Thoracic aortic aneurysm; (k) Venous thromboembolism (all); (l) Deep vein thrombosis; (m) Pulmonary embolism; (n) Peripheral vascular disease; (o) Aortic valve stenosis; (p) Atrial fibrillation; (q) Heart failure; (r) Dilated cardiomyopathy; (s) Endocarditis; (t) Rheumatic heart disease; (u) Chronic kidney disease.**

(i)

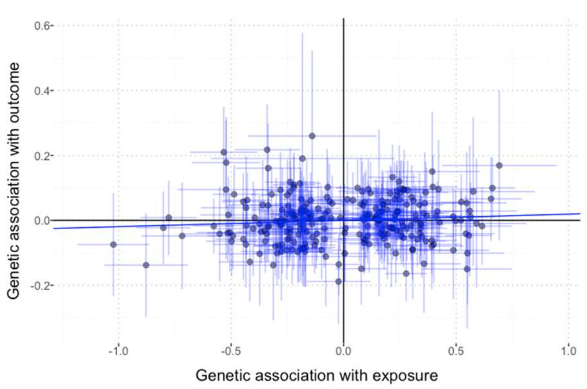


(j)

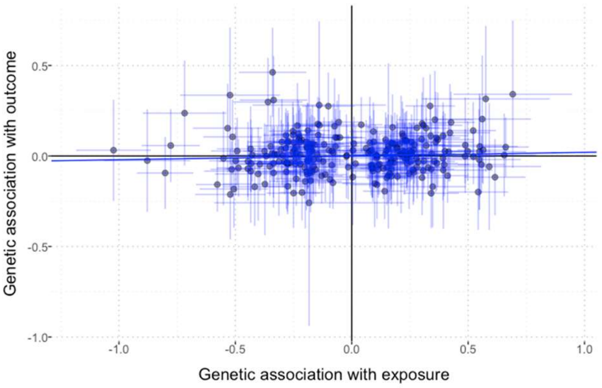


(k)

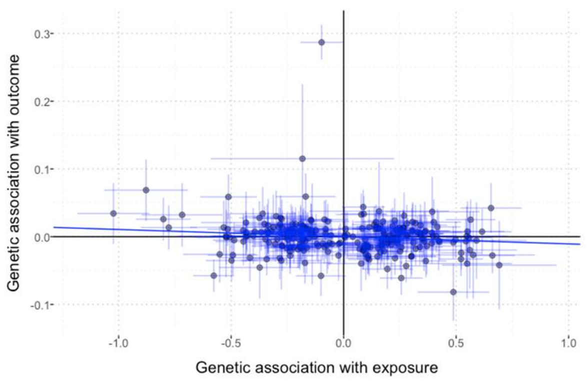


(l)

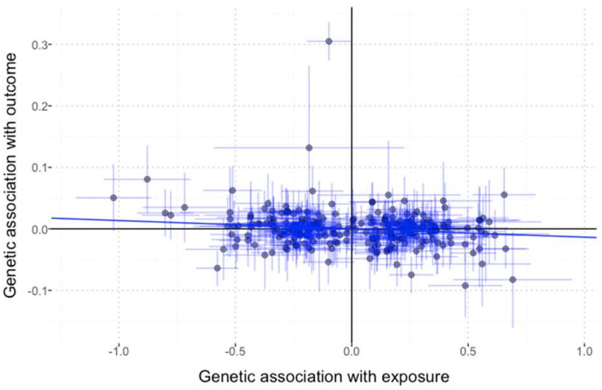


(m)

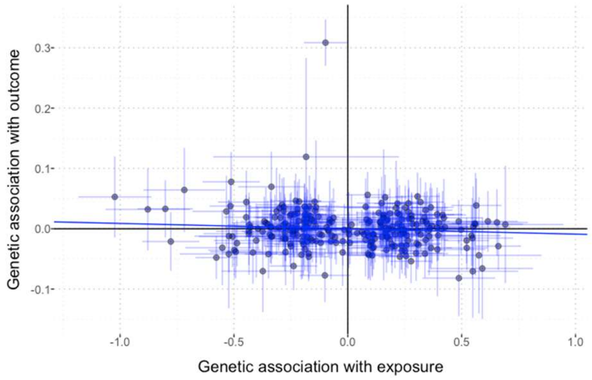


(n)

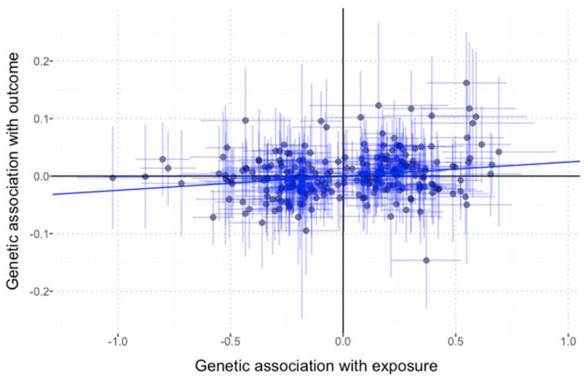


(o)

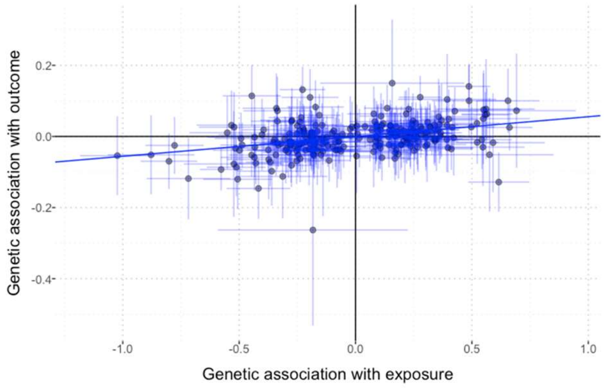


(p)

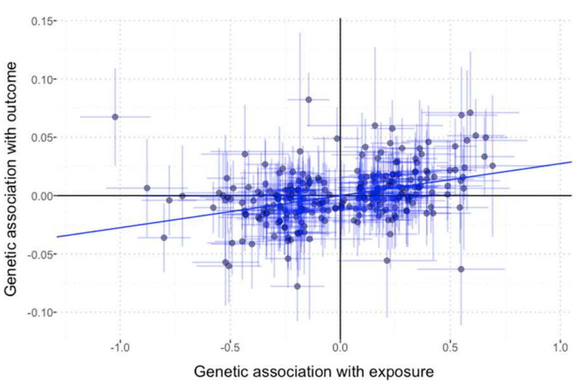


**eFigure 3 (contd.) (a)-(m). Scatter plots of the beta-coefficients for the exposure plotted against the beta-coefficients for the outcome for each of the 256 SNPs in the instrument. Each dot corresponds to a SNP. Lines represent the 95% CI for each SNP's beta-coefficient. Each scatter plot corresponds to one outcome: (a) Coronary artery disease; (b) Ischemic cerebrovascular disease (all); (c) Ischemic stroke; (d) Transient ischemic attack; (e) Hemorrhagic stroke (all); (f) Intracerebral hemorrhage; (g) Subarachnoid hemorrhage; (h) Aortic aneurysm (all); (i) Abdominal aortic aneurysm; (j) Thoracic aortic aneurysm; (k) Venous thromboembolism (all); (l) Deep vein thrombosis; (m) Pulmonary embolism; (n) Peripheral vascular disease; (o) Aortic valve stenosis; (p) Atrial fibrillation; (q) Heart failure; (r) Dilated cardiomyopathy; (s) Endocarditis; (t) Rheumatic heart disease; (u) Chronic kidney disease.**

**(q)**

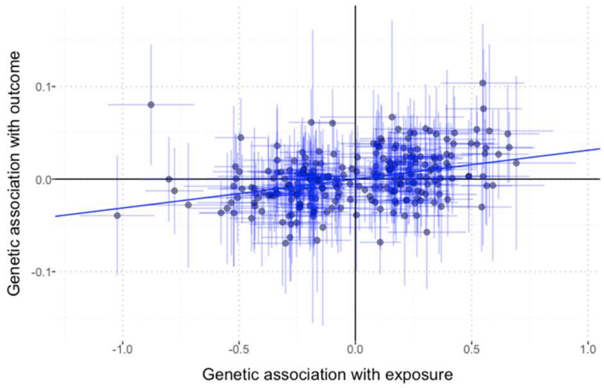


**(r)**

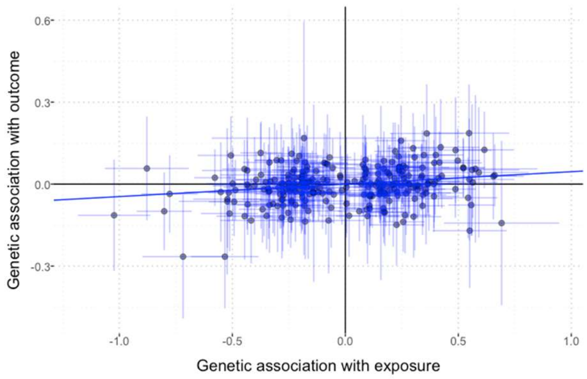


**(s)**

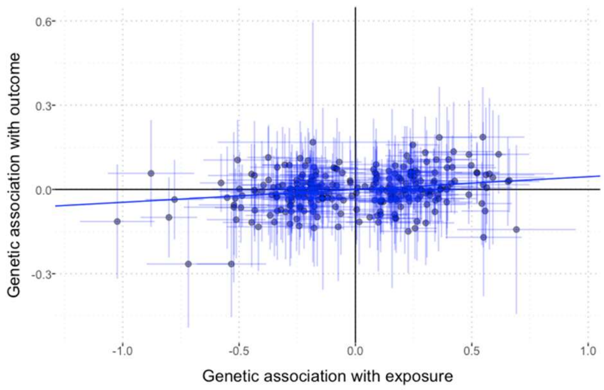


**(t)**

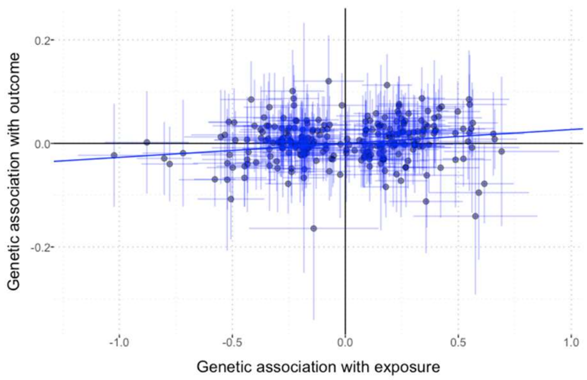


**(u)**

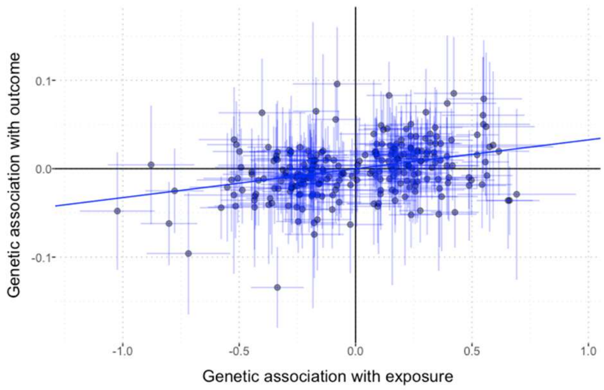


**eFigure 3 (contd.) (a)-(m).** Scatter plots of the beta-coefficients for the exposure plotted against the beta-coefficients for the **outcome for each of the 256 SNPs in the instrument. Each dot corresponds to a SNP. Lines represent the 95% CI for each SNP's beta-coefficient. Each scatter plot corresponds to one outcome: (a) Coronary artery disease; (b) Ischemic cerebrovascular disease (all); (c) Ischemic stroke; (d) Transient ischemic attack; (e) Hemorrhagic stroke (all); (f) Intracerebral hemorrhage; (g) Subarachnoid hemorrhage; (h) Aortic aneurysm (all); (i) Abdominal aortic aneurysm; (j) Thoracic aortic aneurysm; (k) Venous thromboembolism (all); (l) Deep vein thrombosis; (m) Pulmonary embolism; (n) Peripheral vascular disease; (o) Aortic valve stenosis; (p) Atrial fibrillation; (q) Heart failure; (r) Dilated cardiomyopathy; (s) Endocarditis; (t) Rheumatic heart disease; (u) Chronic kidney disease.**

**eTable 1. Summary of disease outcomes considered, definitions, and sources of information within UKBB**

| Outcome name | ICD-9 Diagnosis or death | ICD-10 Diagnosis or death | OPCS procedure code | Self-report[a] |
|---|---|---|---|---|
| **Coronary Artery Disease** | 410.X, 411.X, 412.X, 414.0, 414.8, 414.9 | I21.X, I22.X, I23.X, I24.X, I25.1, I25.2, I25.5, I25.6, I25.8, I25.9 | K40.X, K41.X, K42.X, K43.X, K44.X, K45.X, K46.X, K49.X, K50.1, K50.2, K50.4, K75.X | Non-cancer illness code (20002), Surgical operation code (20004), Health condition diagnosed by doctor (6150) |
| **Ischemic Cerebrovascular Disease (all)** | 434.X, 435.X, 436.X | G45.X, I63.X, I64.X | | Non-cancer illness code (20002) |
| **Ischemic Stroke** | 433.X, 434.X | I63.X, I64.X | | Non-cancer illness code (20002) |
| **Transient Ischemic Attack** | 435.X | G45.X | | Non-cancer illness code (20002) |
| **Hemorrhagic Stroke (all)** | 430.X, 431.X | I60.X, I61.X | | Non-cancer illness code (20002) |
| **Intracerebral Hemorrhage** | 431.X | I61.X | | Non-cancer illness code (20002) |
| **Subarachnoid Hemorrhage** | 430.X | I60.X | | Non-cancer illness code (20002) |
| **Aortic Aneurysm (all)** | 441.X | I71.X | L19.4, L19.5 | Non-cancer illness code (20002), Surgical operation code (20004) |
| **Abdominal Aortic Aneurysm** | 441.3, 441.4 | I71.3, I71.4 | L19.4, L19.5 | Non-cancer illness code (20002) |
| **Thoracic Aortic Aneurysm** | 441.1, 441.2 | I71.1, I71.2 | | Non-cancer illness code (20002) |
| **Venous Thromboembolism (all)** | 415.1, 451.1, 452.X, 453.0, 453.4, 453.9, | I26.X, I80.1, I80.2, I81.X, I82.0 | L90.2 | Non-cancer illness code (20002), Health condition diagnosed by doctor (6152) |
| **Deep Vein Thrombosis** | 451.1 | I80.2 | L90.2 | Non-cancer illness code (20002), Health condition diagnosed by doctor (6152) |
| **Pulmonary Embolism** | 415.1 | I26.X | | Non-cancer illness code (20002), Health condition diagnosed by doctor (6152) |

| Outcome name | ICD-9 Diagnosis or death | ICD-10 Diagnosis or death | OPCS procedure code | Self-report[a] |
|---|---|---|---|---|
| **Peripheral Vascular Disease** | 443.8, 443.9 | I73.8, I73.9 | | Non-cancer illness code (20002) |
| **Aortic Valve Stenosis** | 424.1 | I35.0, I35.2 | | Non-cancer illness code (20002) |
| **Atrial Fibrillation** | 427.3 | I48 | | Non-cancer illness code (20002) |
| **Heart Failure** | 402.01, 402.11, 402.91, 404.01, 404.11, 404.91, 404.03, 404.13, 404.93, 428.X | I11.0, I13.0, I13.2, I50.X | | Non-cancer illness code (20002) |
| **Dilated Cardiomyopathy** | | I42.0 | | |
| **Endocarditis** | 391.1, 421.0, 421.9, 421.1 | I33.X, I38, I39.8, I01.1 | | |
| **Rheumatic Heart Disease** | 391.X, 397.9, 398.0, 394.1, 398.90, 397.1 | I01.X, I02.0, I05.X, I06.X, I07.X, I08.X, I09.X | | |
| **Chronic Kidney Disease** | 585.X | N18.X | | Non-cancer illness code (20002) |

Note that .X means that all sub-codes are matched.
ICD: International Classification of Disease; OPCS: Office of Population Censuses and Surveys Classification of Surgical Operations and Procedures.
[a] Health condition diagnosed by doctor (6150/6152) and Medication for health condition (6177) were self-reported from touchscreen; Non-cancer illness code (20002) and Surgical operation code (20004) were self-reported from interview with trained nurse.

**eTable 2. Details of the 256 single nucleotide polymorphisms (SNPs) from the International Consortium for Blood Pressure (ICBP) genome-wide association study (GWAS) included in the genetic instrument. $\beta_{xj}$ corresponds to the variant-SBP association estimates (beta-coefficients). These variants are taken from Supplementary Table 24 of Evangelou et al, Nat Genet 2018, and represent variants that were published prior to the inclusion of UK Biobank in the ICBP.**

| SNP | Chromosome number | Chromosome position | Effect allele | Other allele | Effect allele frequency | R-squared | F-statistic | $\beta_{xj}$ | $\beta_{xj}$ Standard error | $\beta_{xj}$ P-value |
|---|---|---|---|---|---|---|---|---|---|---|
| rs10057188 | 5 | 77837789 | A | G | 0.4516 | 5.0E-05 | 5.5E-02 | -1.9E-01 | 4.8E-02 | 4.4E-05 |
| rs10059921 | 5 | 87514515 | T | G | 0.0846 | 5.8E-05 | 6.3E-02 | -3.7E-01 | 9.2E-02 | 4.9E-05 |
| rs10077885 | 5 | 114390121 | A | C | 0.498 | 8.2E-05 | 8.9E-02 | -2.5E-01 | 4.8E-02 | 3.5E-07 |
| rs10078021 | 5 | 75038431 | G | T | 0.3744 | 4.8E-06 | 5.3E-03 | 6.2E-02 | 4.9E-02 | 2.1E-01 |
| rs10224002 | 7 | 151415041 | G | A | 0.2814 | 6.1E-05 | 6.7E-02 | 2.4E-01 | 5.3E-02 | 6.0E-06 |
| rs1036477 | 15 | 48914926 | G | A | 0.1044 | 5.8E-05 | 6.3E-02 | -3.4E-01 | 7.4E-02 | 4.1E-06 |
| rs10418305 | 19 | 15278808 | G | C | 0.897 | 3.0E-06 | 3.3E-03 | 7.8E-02 | 7.5E-02 | 3.0E-01 |
| rs1055144 | 7 | 25871109 | T | C | 0.1925 | 9.7E-06 | 1.1E-02 | 1.1E-01 | 5.8E-02 | 6.3E-02 |
| rs1060105 | 12 | 123806219 | T | C | 0.2021 | 8.8E-06 | 9.6E-03 | -1.0E-01 | 5.7E-02 | 7.9E-02 |
| rs1063281 | 2 | 218668732 | T | C | 0.6045 | 7.3E-05 | 8.0E-02 | -2.4E-01 | 4.8E-02 | 7.1E-07 |
| rs10818775 | 9 | 125755571 | T | C | 0.1236 | 4.3E-05 | 4.7E-02 | -2.7E-01 | 7.0E-02 | 8.7E-05 |
| rs10826995 | 10 | 32082658 | C | T | 0.2787 | 8.7E-06 | 9.5E-03 | 9.0E-02 | 5.1E-02 | 8.1E-02 |
| rs10850411 | 12 | 115387796 | C | T | 0.3006 | 1.0E-04 | 1.1E-01 | -3.0E-01 | 4.9E-02 | 8.1E-10 |
| rs10922502 | 1 | 89360158 | G | A | 0.3593 | 6.4E-05 | 7.0E-02 | 2.3E-01 | 4.8E-02 | 2.3E-06 |
| rs10943605 | 6 | 79655477 | A | G | 0.4886 | 4.1E-05 | 4.5E-02 | 1.7E-01 | 4.6E-02 | 1.6E-04 |
| rs11008355 | 10 | 31412561 | C | G | 0.2369 | 7.1E-06 | 7.7E-03 | -8.6E-02 | 5.4E-02 | 1.1E-01 |
| rs11030119 | 11 | 27728102 | A | G | 0.2944 | 3.2E-05 | 3.5E-02 | -1.7E-01 | 5.1E-02 | 8.5E-04 |
| rs110419 | 11 | 8252853 | G | A | 0.5071 | 8.8E-06 | 9.6E-03 | -8.1E-02 | 4.7E-02 | 8.5E-02 |
| rs11067763 | 12 | 116198341 | G | A | 0.1031 | 1.8E-05 | 2.0E-02 | -1.9E-01 | 7.5E-02 | 1.1E-02 |
| rs111245230 | 9 | 113169775 | C | T | 0.0338 | 8.4E-05 | 9.2E-02 | 6.9E-01 | 1.3E-01 | 1.0E-07 |
| rs11128722 | 3 | 14958126 | A | G | 0.5628 | 8.4E-05 | 9.1E-02 | -2.5E-01 | 4.7E-02 | 8.5E-08 |
| rs11154027 | 6 | 121781390 | C | T | 0.5513 | 1.1E-07 | 1.2E-04 | 9.0E-03 | 4.8E-02 | 8.5E-01 |
| rs11191548 | 10 | 104846178 | C | T | 0.0871 | 4.5E-04 | 4.9E-01 | -1.0E+00 | 8.2E-02 | 6.2E-36 |
| rs112184198 | 10 | 102604514 | A | G | 0.1058 | 1.4E-04 | 1.6E-01 | -5.3E-01 | 7.6E-02 | 2.4E-12 |
| rs11229457 | 11 | 58207203 | T | C | 0.2144 | 7.5E-05 | 8.2E-02 | -2.9E-01 | 5.6E-02 | 3.0E-07 |
| rs112557609 | 1 | 56576924 | A | G | 0.3414 | 6.9E-05 | 7.5E-02 | 2.4E-01 | 4.9E-02 | 1.1E-06 |
| rs1126464 | 16 | 89704365 | C | G | 0.245 | 3.4E-05 | 3.7E-02 | 1.8E-01 | 5.7E-02 | 1.1E-03 |
| rs1126930 | 12 | 49399132 | C | G | 0.0343 | 5.9E-05 | 6.4E-02 | 5.8E-01 | 1.4E-01 | 3.9E-05 |
| rs11537751 | 11 | 47587452 | T | C | 0.0521 | 4.1E-05 | 4.5E-02 | 3.9E-01 | 1.1E-01 | 2.6E-04 |
| rs11556924 | 7 | 129663496 | T | C | 0.3713 | 7.8E-05 | 8.5E-02 | -2.5E-01 | 5.0E-02 | 5.9E-07 |
| rs11639856 | 16 | 24788645 | A | T | 0.1902 | 4.3E-05 | 4.7E-02 | -2.3E-01 | 5.8E-02 | 6.8E-05 |
| rs11689667 | 2 | 85491365 | C | T | 0.4536 | 1.9E-05 | 2.1E-02 | -1.2E-01 | 4.7E-02 | 1.1E-02 |
| rs11690961 | 2 | 46363336 | C | A | 0.116 | 2.9E-07 | 3.1E-04 | -2.3E-02 | 7.2E-02 | 7.5E-01 |
| rs11701033 | 21 | 33788341 | G | C | 0.1831 | 4.9E-05 | 5.3E-02 | 2.5E-01 | 5.9E-02 | 3.2E-05 |
| rs1173771 | 5 | 32815028 | G | A | 0.6024 | 3.5E-04 | 3.8E-01 | 5.2E-01 | 4.7E-02 | 6.0E-29 |
| rs11953630 | 5 | 157845402 | T | C | 0.3694 | 2.5E-04 | 2.7E-01 | -4.5E-01 | 5.0E-02 | 5.2E-19 |
| rs11977526 | 7 | 46008110 | A | G | 0.3993 | 8.3E-05 | 9.0E-02 | -2.5E-01 | 4.8E-02 | 1.6E-07 |
| rs12374077 | 3 | 185317674 | C | G | 0.3452 | 3.5E-05 | 3.8E-02 | 1.7E-01 | 4.8E-02 | 4.6E-04 |
| rs12405515 | 1 | 172357441 | T | G | 0.5741 | 3.9E-05 | 4.2E-02 | -1.7E-01 | 4.6E-02 | 2.0E-04 |
| rs12408022 | 1 | 217718789 | T | C | 0.2611 | 6.5E-05 | 7.1E-02 | 2.5E-01 | 5.3E-02 | 2.4E-06 |
| rs12521868 | 5 | 131784393 | T | G | 0.4156 | 2.0E-05 | 2.2E-02 | -1.2E-01 | 4.8E-02 | 9.7E-03 |
| rs12579720 | 12 | 20173764 | G | C | 0.7621 | 8.9E-05 | 9.7E-02 | 3.0E-01 | 5.4E-02 | 2.5E-08 |
| rs12627651 | 21 | 44760603 | A | G | 0.2944 | 1.9E-04 | 2.1E-01 | 4.2E-01 | 5.3E-02 | 3.3E-15 |
| rs12628032 | 22 | 19967980 | T | C | 0.3152 | 4.5E-05 | 4.9E-02 | 2.0E-01 | 5.0E-02 | 7.7E-05 |
| rs1275988 | 2 | 26914364 | T | C | 0.6055 | 3.4E-04 | 3.7E-01 | -5.2E-01 | 4.7E-02 | 1.8E-28 |
| rs12906962 | 15 | 95312071 | C | T | 0.3293 | 8.4E-05 | 9.2E-02 | 2.7E-01 | 4.9E-02 | 6.4E-08 |
| rs12921187 | 16 | 4943019 | G | T | 0.5726 | 7.7E-05 | 8.4E-02 | 2.4E-01 | 4.6E-02 | 1.3E-07 |
| rs12940887 | 17 | 47402807 | T | C | 0.3727 | 8.9E-05 | 9.7E-02 | 2.7E-01 | 4.7E-02 | 1.4E-08 |
| rs12941318 | 17 | 1333598 | C | T | 0.5018 | 5.8E-05 | 6.3E-02 | 2.1E-01 | 4.8E-02 | 1.7E-05 |
| rs12946454 | 17 | 43208121 | T | A | 0.261 | 1.1E-04 | 1.2E-01 | 3.2E-01 | 5.2E-02 | 7.3E-10 |
| rs12958173 | 18 | 42141977 | C | A | 0.7 | 1.4E-04 | 1.5E-01 | -3.5E-01 | 5.0E-02 | 1.2E-12 |
| rs13082711 | 3 | 27537909 | C | T | 0.231 | 7.5E-05 | 8.2E-02 | 2.8E-01 | 5.5E-02 | 2.8E-07 |
| rs13107325 | 4 | 103188709 | T | C | 0.0722 | 2.8E-04 | 3.0E-01 | -8.8E-01 | 9.5E-02 | 1.8E-20 |
| rs13112725 | 4 | 106911742 | C | G | 0.7682 | 1.5E-04 | 1.6E-01 | 4.0E-01 | 5.6E-02 | 1.0E-12 |

| SNP | Chromosome number | Chromosome position | Effect allele | Other allele | Effect allele frequency | R-squared | F-statistic | $\beta_{xj}$ | $\beta_{xj}$ Standard error | $\beta_{xj}$ P-value |
|---|---|---|---|---|---|---|---|---|---|---|
| rs13139571 | 4 | 156645513 | A | C | 0.2393 | 8.6E-05 | 9.3E-02 | -3.0E-01 | 5.4E-02 | 3.7E-08 |
| rs13205180 | 6 | 51832494 | T | C | 0.4808 | 2.3E-06 | 2.5E-03 | 4.1E-02 | 4.6E-02 | 3.8E-01 |
| rs13209747 | 6 | 127115454 | T | C | 0.4465 | 2.2E-04 | 2.5E-01 | 4.1E-01 | 4.7E-02 | 9.8E-19 |
| rs1322639 | 6 | 169587103 | A | G | 0.7711 | 7.5E-06 | 8.2E-03 | 8.9E-02 | 5.6E-02 | 1.1E-01 |
| rs13238550 | 7 | 131059056 | A | G | 0.3909 | 3.7E-05 | 4.0E-02 | 1.7E-01 | 4.7E-02 | 3.3E-04 |
| rs1327235 | 20 | 10969030 | G | A | 0.4631 | 1.7E-04 | 1.8E-01 | 3.5E-01 | 4.5E-02 | 3.2E-15 |
| rs13303 | 3 | 52558008 | C | T | 0.5697 | 1.6E-05 | 1.7E-02 | 1.1E-01 | 4.7E-02 | 2.1E-02 |
| rs13333226 | 16 | 20365654 | G | A | 0.186 | 9.2E-05 | 1.0E-01 | -3.4E-01 | 5.8E-02 | 7.0E-09 |
| rs13359291 | 5 | 122476457 | A | G | 0.1654 | 1.2E-04 | 1.3E-01 | 4.0E-01 | 6.2E-02 | 1.1E-10 |
| rs13420463 | 2 | 37517566 | G | A | 0.2225 | 7.0E-05 | 7.7E-02 | -2.8E-01 | 5.6E-02 | 7.2E-07 |
| rs1344653 | 2 | 19730845 | G | A | 0.5039 | 3.3E-05 | 3.6E-02 | 1.6E-01 | 4.6E-02 | 5.8E-04 |
| rs1378942 | 15 | 75077367 | A | C | 0.6563 | 2.9E-04 | 3.1E-01 | -4.9E-01 | 4.8E-02 | 5.0E-24 |
| rs143112823 | 3 | 154707967 | A | G | 0.076 | 6.1E-05 | 6.6E-02 | -4.0E-01 | 9.5E-02 | 2.3E-05 |
| rs1438896 | 2 | 145646072 | C | T | 0.7001 | 5.8E-05 | 6.4E-02 | -2.3E-01 | 5.0E-02 | 5.4E-06 |
| rs1446468 | 2 | 164963486 | C | T | 0.5488 | 3.2E-04 | 3.4E-01 | 4.9E-01 | 4.7E-02 | 2.3E-25 |
| rs1449544 | 8 | 76591880 | C | A | 0.4584 | 6.5E-05 | 7.1E-02 | -2.2E-01 | 4.6E-02 | 1.4E-06 |
| rs1458038 | 4 | 81164723 | T | C | 0.2971 | 4.9E-04 | 5.4E-01 | 6.6E-01 | 5.1E-02 | 6.1E-39 |
| rs1475130 | 14 | 100225144 | C | T | 0.6571 | 3.2E-05 | 3.5E-02 | 1.6E-01 | 4.9E-02 | 7.4E-04 |
| rs147696085 | 1 | 51021867 | A | G | 0.1011 | 2.7E-06 | 2.9E-03 | -7.4E-02 | 8.0E-02 | 3.5E-01 |
| rs1530440 | 10 | 63524591 | T | C | 0.1874 | 2.0E-04 | 2.2E-01 | -5.0E-01 | 5.9E-02 | 2.0E-17 |
| rs1563788 | 6 | 43308363 | T | C | 0.2937 | 1.0E-04 | 1.1E-01 | 3.1E-01 | 5.0E-02 | 9.8E-10 |
| rs167479 | 19 | 11526765 | T | G | 0.4732 | 2.5E-04 | 2.7E-01 | -4.3E-01 | 5.6E-02 | 2.5E-14 |
| rs16823124 | 2 | 183224127 | A | G | 0.303 | 6.1E-05 | 6.7E-02 | 2.3E-01 | 5.0E-02 | 3.1E-06 |
| rs16851397 | 3 | 141134818 | G | A | 0.0467 | 8.3E-05 | 9.1E-02 | 5.9E-01 | 1.1E-01 | 1.9E-07 |
| rs17030613 | 1 | 113190807 | C | A | 0.2191 | 1.4E-04 | 1.5E-01 | 3.9E-01 | 5.6E-02 | 2.5E-12 |
| rs17080102 | 6 | 151004770 | C | G | 0.0677 | 1.7E-04 | 1.9E-01 | -7.2E-01 | 9.2E-02 | 4.9E-15 |
| rs17249754 | 12 | 90060586 | A | G | 0.1637 | 4.7E-04 | 5.2E-01 | -8.0E-01 | 6.2E-02 | 2.2E-38 |
| rs17367504 | 1 | 11862778 | G | A | 0.1556 | 4.3E-04 | 4.7E-01 | -7.8E-01 | 6.4E-02 | 4.8E-34 |
| rs17477177 | 7 | 106411858 | C | T | 0.2094 | 2.8E-04 | 3.1E-01 | 5.6E-01 | 5.6E-02 | 1.6E-23 |
| rs17608766 | 17 | 45013271 | C | T | 0.1433 | 2.5E-04 | 2.7E-01 | 6.2E-01 | 6.7E-02 | 3.9E-20 |
| rs17638167 | 19 | 11584818 | T | C | 0.047 | 6.6E-05 | 7.2E-02 | -5.2E-01 | 1.1E-01 | 1.8E-06 |
| rs1799945 | 6 | 26091179 | G | C | 0.1479 | 2.0E-04 | 2.2E-01 | 5.4E-01 | 6.6E-02 | 1.3E-16 |
| rs1813353 | 10 | 18707448 | C | T | 0.3411 | 2.3E-04 | 2.5E-01 | -4.3E-01 | 4.8E-02 | 3.0E-19 |
| rs1876487 | 2 | 73114352 | C | A | 0.7122 | 7.5E-06 | 8.2E-03 | 8.3E-02 | 5.5E-02 | 1.3E-01 |
| rs1925153 | 6 | 56102780 | T | C | 0.4401 | 3.5E-06 | 3.8E-03 | -5.1E-02 | 4.9E-02 | 3.0E-01 |
| rs1953126 | 9 | 123640500 | C | T | 0.6453 | 4.2E-05 | 4.5E-02 | -1.8E-01 | 4.8E-02 | 1.2E-04 |
| rs1975487 | 2 | 55809054 | G | A | 0.5124 | 6.5E-05 | 7.1E-02 | 2.2E-01 | 4.7E-02 | 2.9E-06 |
| rs2004776 | 1 | 230848702 | T | C | 0.2381 | 1.1E-04 | 1.2E-01 | 3.4E-01 | 5.4E-02 | 5.3E-10 |
| rs2014912 | 4 | 86715670 | C | T | 0.8485 | 1.8E-04 | 2.0E-01 | -5.1E-01 | 6.4E-02 | 1.8E-15 |
| rs2034618 | 15 | 83799632 | T | C | 0.2231 | 2.1E-07 | 2.3E-04 | -1.5E-02 | 5.5E-02 | 7.8E-01 |
| rs2071518 | 8 | 120435812 | T | C | 0.2571 | 4.9E-05 | 5.3E-02 | 2.2E-01 | 5.2E-02 | 3.1E-05 |
| rs2076328 | 1 | 1687482 | T | G | 0.4748 | 1.1E-04 | 1.2E-01 | -2.8E-01 | 5.0E-02 | 1.4E-08 |
| rs2107595 | 7 | 19049388 | A | G | 0.166 | 6.8E-05 | 7.4E-02 | 3.0E-01 | 6.2E-02 | 1.2E-06 |
| rs2157597 | 1 | 169201567 | T | C | 0.3468 | 8.9E-06 | 9.7E-03 | -8.6E-02 | 4.9E-02 | 8.1E-02 |
| rs2240736 | 17 | 59485393 | T | C | 0.7328 | 1.9E-04 | 2.1E-01 | 4.3E-01 | 5.3E-02 | 4.5E-16 |
| rs2246438 | 10 | 45273079 | A | G | 0.2779 | 5.1E-07 | 5.5E-04 | -2.2E-02 | 5.1E-02 | 6.7E-01 |
| rs2282978 | 7 | 92264410 | C | T | 0.342 | 6.0E-06 | 6.5E-03 | -7.0E-02 | 4.9E-02 | 1.5E-01 |
| rs2289125 | 11 | 89224453 | C | A | 0.7697 | 3.6E-05 | 3.9E-02 | 1.9E-01 | 5.8E-02 | 8.5E-04 |
| rs2291435 | 4 | 38387395 | T | C | 0.5248 | 7.8E-05 | 8.6E-02 | -2.4E-01 | 4.6E-02 | 1.7E-07 |
| rs2304130 | 19 | 19789528 | G | A | 0.0833 | 3.9E-05 | 4.2E-02 | 3.1E-01 | 8.6E-02 | 3.6E-04 |
| rs2306374 | 3 | 138119952 | C | T | 0.1619 | 3.7E-05 | 4.1E-02 | 2.3E-01 | 6.3E-02 | 3.0E-04 |
| rs2404715 | 1 | 57008778 | T | C | 0.0925 | 1.8E-05 | 2.0E-02 | -2.0E-01 | 8.0E-02 | 1.2E-02 |
| rs2467099 | 17 | 73949045 | T | C | 0.2204 | 3.4E-05 | 3.7E-02 | -1.9E-01 | 5.5E-02 | 4.8E-04 |
| rs2493292 | 1 | 3328659 | T | C | 0.1501 | 4.9E-05 | 5.4E-02 | 2.7E-01 | 6.8E-02 | 7.8E-05 |
| rs2521501 | 15 | 91437388 | T | A | 0.3301 | 3.7E-04 | 4.0E-01 | 5.6E-01 | 5.2E-02 | 4.8E-27 |
| rs2579519 | 2 | 96675166 | C | T | 0.3949 | 3.7E-08 | 4.1E-05 | 5.4E-03 | 4.8E-02 | 9.1E-01 |
| rs2645466 | 17 | 57853214 | C | A | 0.3018 | 4.4E-05 | 4.8E-02 | 2.0E-01 | 4.9E-02 | 7.1E-05 |
| rs2759308 | 15 | 81016227 | A | G | 0.4758 | 9.0E-05 | 9.8E-02 | 2.6E-01 | 4.6E-02 | 1.8E-08 |
| rs2761436 | 1 | 207919748 | T | C | 0.5276 | 5.0E-05 | 5.4E-02 | 1.9E-01 | 4.6E-02 | 2.9E-05 |
| rs2782980 | 10 | 115781527 | C | T | 0.7096 | 1.3E-04 | 1.4E-01 | 3.4E-01 | 5.2E-02 | 7.6E-11 |

| SNP | Chromosome number | Chromosome position | Effect allele | Other allele | Effect allele frequency | R-squared | F-statistic | $\beta_{xj}$ | $\beta_{xj}$ Standard error | $\beta_{xj}$ P-value |
|---|---|---|---|---|---|---|---|---|---|---|
| rs28427409 | 17 | 6473882 | T | C | 0.4133 | 3.5E-05 | 3.8E-02 | 1.6E-01 | 4.6E-02 | 4.0E-04 |
| rs2898290 | 8 | 11433909 | C | T | 0.5165 | 1.6E-04 | 1.7E-01 | -3.4E-01 | 4.7E-02 | 2.1E-13 |
| rs2932538 | 1 | 113216543 | G | A | 0.7414 | 1.2E-04 | 1.3E-01 | 3.4E-01 | 5.3E-02 | 7.2E-11 |
| rs2969070 | 7 | 2512545 | A | G | 0.6369 | 9.4E-05 | 1.0E-01 | -2.8E-01 | 4.8E-02 | 8.4E-09 |
| rs2972146 | 2 | 227100698 | T | G | 0.6362 | 7.7E-05 | 8.4E-02 | 2.5E-01 | 4.8E-02 | 1.8E-07 |
| rs2978098 | 8 | 101676675 | C | A | 0.442 | 2.9E-05 | 3.2E-02 | -1.5E-01 | 4.7E-02 | 1.7E-03 |
| rs2978456 | 8 | 42324765 | C | T | 0.4442 | 1.5E-05 | 1.7E-02 | 1.1E-01 | 5.0E-02 | 3.2E-02 |
| rs3184504 | 12 | 111884608 | C | T | 0.5244 | 4.5E-04 | 4.9E-01 | -5.8E-01 | 4.7E-02 | 2.6E-35 |
| rs33063 | 16 | 69640217 | G | A | 0.856 | 2.1E-05 | 2.3E-02 | -1.8E-01 | 6.5E-02 | 6.0E-03 |
| rs34591516 | 8 | 142367087 | T | C | 0.0533 | 8.5E-05 | 9.3E-02 | 5.6E-01 | 1.1E-01 | 1.5E-07 |
| rs347591 | 3 | 11290122 | T | G | 0.6625 | 9.7E-05 | 1.1E-01 | 2.8E-01 | 4.9E-02 | 6.3E-09 |
| rs34872471 | 10 | 114754071 | C | T | 0.2936 | 3.6E-05 | 3.9E-02 | 1.8E-01 | 5.1E-02 | 4.2E-04 |
| rs35261357 | 16 | 75444572 | T | C | 0.5698 | 1.0E-04 | 1.1E-01 | 2.8E-01 | 4.7E-02 | 2.7E-09 |
| rs35410524 | 6 | 96885405 | T | C | 0.1917 | 7.5E-05 | 8.2E-02 | 3.0E-01 | 5.9E-02 | 3.4E-07 |
| rs35444 | 12 | 115552437 | G | A | 0.3921 | 1.4E-04 | 1.5E-01 | -3.3E-01 | 4.7E-02 | 1.8E-12 |
| rs35783704 | 8 | 105966258 | A | G | 0.1092 | 1.4E-04 | 1.6E-01 | -5.2E-01 | 7.7E-02 | 1.5E-11 |
| rs36010659 | 18 | 48283949 | C | T | 0.1391 | 2.4E-05 | 2.7E-02 | -2.0E-01 | 6.6E-02 | 3.0E-03 |
| rs36022378 | 3 | 49913705 | C | T | 0.1926 | 2.0E-05 | 2.2E-02 | 1.6E-01 | 6.0E-02 | 9.3E-03 |
| rs3741378 | 11 | 65408937 | T | C | 0.1328 | 1.1E-04 | 1.2E-01 | -4.2E-01 | 7.0E-02 | 2.2E-09 |
| rs3771371 | 2 | 71627539 | T | C | 0.5658 | 4.3E-05 | 4.7E-02 | -1.8E-01 | 4.6E-02 | 9.2E-05 |
| rs381815 | 11 | 16902268 | T | C | 0.2705 | 1.4E-04 | 1.5E-01 | 3.6E-01 | 5.2E-02 | 1.5E-12 |
| rs3820068 | 1 | 15798197 | G | A | 0.2023 | 9.8E-05 | 1.1E-01 | -3.4E-01 | 6.0E-02 | 1.7E-08 |
| rs3918226 | 7 | 150690176 | T | C | 0.0811 | 1.2E-04 | 1.3E-01 | 5.5E-01 | 9.1E-02 | 1.8E-09 |
| rs409558 | 6 | 31708147 | C | T | 0.1712 | 1.5E-04 | 1.7E-01 | -4.5E-01 | 6.4E-02 | 2.6E-12 |
| rs419076 | 3 | 169100886 | C | T | 0.5265 | 2.1E-04 | 2.3E-01 | -3.9E-01 | 4.6E-02 | 5.4E-18 |
| rs4245739 | 1 | 204518842 | A | C | 0.7326 | 3.0E-05 | 3.3E-02 | 1.7E-01 | 5.3E-02 | 1.2E-03 |
| rs4247374 | 19 | 7252756 | T | C | 0.1355 | 1.6E-04 | 1.8E-01 | -5.1E-01 | 7.5E-02 | 1.8E-11 |
| rs4292285 | 4 | 145271954 | A | T | 0.3994 | 2.1E-05 | 2.3E-02 | -1.3E-01 | 4.7E-02 | 5.8E-03 |
| rs4308 | 17 | 61559625 | G | A | 0.6178 | 9.4E-05 | 1.0E-01 | -2.7E-01 | 4.8E-02 | 1.1E-08 |
| rs4364717 | 9 | 21801530 | G | A | 0.4534 | 1.5E-05 | 1.6E-02 | 1.1E-01 | 4.6E-02 | 1.9E-02 |
| rs4373814 | 10 | 18419972 | C | G | 0.441 | 8.4E-05 | 9.2E-02 | 2.5E-01 | 4.7E-02 | 6.2E-08 |
| rs4387287 | 10 | 105677897 | C | A | 0.8262 | 2.9E-05 | 3.2E-02 | -2.0E-01 | 6.2E-02 | 1.8E-03 |
| rs4454254 | 8 | 141060027 | A | G | 0.6346 | 6.1E-05 | 6.7E-02 | -2.2E-01 | 4.8E-02 | 3.2E-06 |
| rs4494250 | 10 | 96563757 | A | G | 0.3611 | 8.1E-05 | 8.9E-02 | 2.6E-01 | 4.9E-02 | 1.3E-07 |
| rs449789 | 6 | 159699125 | G | C | 0.8652 | 1.8E-05 | 1.9E-02 | -1.7E-01 | 6.8E-02 | 1.3E-02 |
| rs452036 | 14 | 23865885 | A | G | 0.3492 | 2.5E-05 | 2.8E-02 | -1.4E-01 | 4.8E-02 | 2.7E-03 |
| rs470113 | 22 | 40729614 | G | A | 0.188 | 5.0E-06 | 5.5E-03 | 7.8E-02 | 5.8E-02 | 1.8E-01 |
| rs4728142 | 7 | 128573967 | A | G | 0.4383 | 6.1E-05 | 6.7E-02 | -2.2E-01 | 4.7E-02 | 3.9E-06 |
| rs4757391 | 11 | 16302939 | T | C | 0.8013 | 2.1E-04 | 2.3E-01 | -4.9E-01 | 5.7E-02 | 8.3E-18 |
| rs4823006 | 22 | 29451671 | G | A | 0.4384 | 2.6E-05 | 2.9E-02 | -1.4E-01 | 4.6E-02 | 2.0E-03 |
| rs4952611 | 2 | 40567743 | T | C | 0.5834 | 5.9E-05 | 6.5E-02 | -2.1E-01 | 4.9E-02 | 1.2E-05 |
| rs5219 | 11 | 17409572 | C | T | 0.6245 | 1.3E-04 | 1.4E-01 | -3.2E-01 | 4.7E-02 | 1.1E-11 |
| rs55701159 | 2 | 25139596 | G | T | 0.113 | 4.8E-05 | 5.3E-02 | -3.0E-01 | 7.4E-02 | 5.3E-05 |
| rs55780018 | 2 | 208526140 | C | T | 0.452 | 1.4E-04 | 1.6E-01 | 3.3E-01 | 4.9E-02 | 1.9E-11 |
| rs57927100 | 17 | 75317300 | G | C | 0.259 | 1.2E-04 | 1.4E-01 | -3.5E-01 | 5.4E-02 | 8.4E-11 |
| rs6015450 | 20 | 57751117 | G | A | 0.1287 | 2.6E-04 | 2.8E-01 | 6.6E-01 | 6.9E-02 | 2.4E-21 |
| rs60199046 | 1 | 59663341 | G | A | 0.2868 | 3.5E-05 | 3.8E-02 | -1.8E-01 | 5.1E-02 | 4.3E-04 |
| rs6031435 | 20 | 42797358 | G | A | 0.4612 | 6.9E-05 | 7.5E-02 | 2.3E-01 | 4.6E-02 | 6.7E-07 |
| rs6060114 | 20 | 30169673 | C | T | 0.162 | 5.8E-05 | 6.3E-02 | -2.8E-01 | 6.2E-02 | 5.9E-06 |
| rs6081613 | 20 | 19465907 | A | G | 0.2722 | 2.0E-05 | 2.1E-02 | 1.4E-01 | 5.1E-02 | 7.3E-03 |
| rs6095241 | 20 | 47308798 | A | G | 0.4363 | 4.7E-05 | 5.1E-02 | -1.9E-01 | 4.5E-02 | 3.3E-05 |
| rs6108168 | 20 | 8626271 | A | C | 0.2541 | 4.2E-05 | 4.6E-02 | -2.0E-01 | 5.2E-02 | 8.8E-05 |
| rs62011052 | 15 | 79156983 | C | T | 0.1497 | 1.9E-07 | 2.0E-04 | 1.7E-02 | 6.4E-02 | 8.0E-01 |
| rs62104477 | 19 | 30294991 | T | G | 0.3313 | 9.0E-06 | 9.8E-03 | 8.7E-02 | 4.9E-02 | 7.4E-02 |
| rs62270945 | 3 | 128201889 | T | C | 0.0302 | 3.9E-06 | 4.3E-03 | 1.6E-01 | 1.6E-01 | 3.2E-01 |
| rs62524579 | 8 | 144060955 | A | G | 0.5307 | 4.4E-05 | 4.8E-02 | -1.8E-01 | 5.3E-02 | 6.9E-04 |
| rs6271 | 9 | 136522274 | T | C | 0.0725 | 6.8E-05 | 7.4E-02 | -4.3E-01 | 1.0E-01 | 2.1E-05 |
| rs633185 | 11 | 100593538 | C | G | 0.7096 | 2.7E-04 | 3.0E-01 | 5.0E-01 | 5.1E-02 | 9.8E-23 |
| rs6429422 | 1 | 243472801 | G | T | 0.3289 | 2.4E-05 | 2.7E-02 | 1.4E-01 | 4.9E-02 | 3.6E-03 |
| rs6487543 | 12 | 26438189 | A | G | 0.7655 | 3.0E-05 | 3.2E-02 | 1.8E-01 | 5.6E-02 | 1.9E-03 |

| SNP | Chromosome number | Chromosome position | Effect allele | Other allele | Effect allele frequency | R-squared | F-statistic | $\beta_{xj}$ | $\beta_{xj}$ Standard error | $\beta_{xj}$ P-value |
|---|---|---|---|---|---|---|---|---|---|---|
| rs6557876 | 8 | 25900675 | T | C | 0.2511 | 1.4E-04 | 1.5E-01 | -3.7E-01 | 5.3E-02 | 6.0E-12 |
| rs6595838 | 5 | 127868199 | A | G | 0.2891 | 6.2E-05 | 6.7E-02 | 2.4E-01 | 5.1E-02 | 3.1E-06 |
| rs661348 | 11 | 1905292 | C | T | 0.4368 | 1.5E-04 | 1.7E-01 | 3.4E-01 | 5.0E-02 | 9.6E-12 |
| rs6686889 | 1 | 25030470 | T | C | 0.2581 | 7.5E-06 | 8.2E-03 | 8.5E-02 | 5.3E-02 | 1.1E-01 |
| rs66887589 | 4 | 120509279 | C | T | 0.4814 | 4.4E-05 | 4.8E-02 | 1.8E-01 | 4.6E-02 | 9.0E-05 |
| rs67330701 | 11 | 69079707 | T | C | 0.0958 | 3.6E-05 | 3.9E-02 | -2.8E-01 | 9.1E-02 | 2.4E-03 |
| rs6783086 | 3 | 133959552 | T | C | 0.409 | 8.6E-05 | 9.4E-02 | 2.6E-01 | 4.7E-02 | 3.9E-08 |
| rs6797587 | 3 | 48197614 | G | A | 0.6797 | 9.2E-05 | 1.0E-01 | 2.8E-01 | 5.0E-02 | 1.5E-08 |
| rs6825911 | 4 | 111381638 | T | C | 0.787 | 5.1E-05 | 5.5E-02 | -2.4E-01 | 5.8E-02 | 3.7E-05 |
| rs687621 | 9 | 136137065 | G | A | 0.3392 | 1.2E-05 | 1.3E-02 | -9.8E-02 | 4.8E-02 | 4.3E-02 |
| rs6891344 | 5 | 123136656 | G | A | 0.1856 | 6.5E-05 | 7.1E-02 | -2.8E-01 | 6.0E-02 | 2.2E-06 |
| rs6911827 | 6 | 22130601 | T | C | 0.4623 | 3.1E-05 | 3.4E-02 | 1.5E-01 | 4.7E-02 | 1.3E-03 |
| rs6969780 | 7 | 27159136 | C | G | 0.0961 | 6.4E-05 | 7.0E-02 | 3.7E-01 | 7.9E-02 | 3.1E-06 |
| rs709209 | 1 | 6278414 | G | A | 0.3421 | 1.1E-05 | 1.2E-02 | -9.4E-02 | 5.3E-02 | 7.6E-02 |
| rs7103648 | 11 | 47461783 | G | A | 0.3844 | 1.2E-04 | 1.3E-01 | 3.1E-01 | 4.7E-02 | 6.2E-11 |
| rs7126805 | 11 | 828916 | A | G | 0.7292 | 1.5E-05 | 1.7E-02 | 1.2E-01 | 5.6E-02 | 3.1E-02 |
| rs7129220 | 11 | 10350538 | A | G | 0.1233 | 8.9E-05 | 9.7E-02 | 3.9E-01 | 7.2E-02 | 6.3E-08 |
| rs7178615 | 15 | 66869072 | G | A | 0.6085 | 2.6E-05 | 2.8E-02 | 1.4E-01 | 4.7E-02 | 2.8E-03 |
| rs7236548 | 18 | 43097750 | A | C | 0.1857 | 5.7E-05 | 6.2E-02 | 2.6E-01 | 5.8E-02 | 5.5E-06 |
| rs7248104 | 19 | 7224431 | A | G | 0.4085 | 3.3E-05 | 3.6E-02 | -1.6E-01 | 4.6E-02 | 6.0E-04 |
| rs7255 | 2 | 20878820 | C | T | 0.5345 | 6.7E-06 | 7.3E-03 | 7.1E-02 | 4.7E-02 | 1.3E-01 |
| rs72765298 | 9 | 127900996 | C | T | 0.1179 | 6.8E-05 | 7.4E-02 | 3.5E-01 | 7.3E-02 | 1.7E-06 |
| rs72799341 | 16 | 30936743 | A | G | 0.2392 | 8.8E-06 | 9.6E-03 | 9.5E-02 | 5.4E-02 | 7.9E-02 |
| rs72812846 | 5 | 173377636 | A | T | 0.278 | 6.1E-05 | 6.7E-02 | -2.4E-01 | 5.3E-02 | 8.2E-06 |
| rs7297416 | 12 | 54443090 | C | A | 0.3133 | 9.2E-05 | 1.0E-01 | -2.8E-01 | 5.0E-02 | 1.8E-08 |
| rs7302981 | 12 | 50537815 | G | A | 0.6131 | 1.8E-04 | 2.0E-01 | -3.7E-01 | 4.6E-02 | 4.4E-16 |
| rs73030266 | 6 | 166179459 | T | A | 0.0674 | 3.3E-05 | 3.6E-02 | -3.1E-01 | 9.8E-02 | 1.4E-03 |
| rs73091767 | 1 | 227250775 | C | T | 0.2654 | 8.6E-06 | 9.4E-03 | 9.1E-02 | 5.2E-02 | 7.9E-02 |
| rs73099903 | 12 | 53440779 | T | C | 0.0794 | 7.0E-05 | 7.6E-02 | 4.2E-01 | 8.8E-02 | 1.6E-06 |
| rs73161324 | 22 | 42038786 | T | C | 0.0591 | 1.3E-05 | 1.5E-02 | 2.1E-01 | 1.2E-01 | 6.7E-02 |
| rs740406 | 19 | 2232221 | G | A | 0.0631 | 9.6E-05 | 1.0E-01 | 5.5E-01 | 1.0E-01 | 6.9E-08 |
| rs7406910 | 17 | 46688256 | C | T | 0.9107 | 1.0E-04 | 1.1E-01 | 4.9E-01 | 8.1E-02 | 1.9E-09 |
| rs740698 | 17 | 60767151 | T | C | 0.5726 | 2.8E-05 | 3.0E-02 | -1.5E-01 | 4.8E-02 | 2.2E-03 |
| rs745821 | 18 | 48142854 | G | T | 0.2509 | 3.2E-05 | 3.5E-02 | -1.8E-01 | 5.3E-02 | 7.5E-04 |
| rs7480089 | 11 | 45207851 | A | G | 0.1222 | 3.1E-05 | 3.4E-02 | -2.3E-01 | 7.2E-02 | 1.1E-03 |
| rs7500448 | 16 | 83045790 | G | A | 0.2531 | 5.1E-05 | 5.6E-02 | -2.3E-01 | 5.3E-02 | 2.4E-05 |
| rs7515635 | 1 | 42408070 | C | T | 0.5316 | 7.6E-05 | 8.3E-02 | -2.4E-01 | 4.6E-02 | 2.7E-07 |
| rs751984 | 11 | 61278246 | C | T | 0.1211 | 1.1E-04 | 1.2E-01 | -4.3E-01 | 7.4E-02 | 4.1E-09 |
| rs7562 | 2 | 28635740 | C | T | 0.4703 | 3.2E-05 | 3.5E-02 | -1.6E-01 | 4.7E-02 | 9.3E-04 |
| rs7592578 | 2 | 191439591 | G | T | 0.7926 | 1.0E-04 | 1.1E-01 | 3.4E-01 | 6.0E-02 | 1.5E-08 |
| rs76206723 | 7 | 40447971 | A | G | 0.1097 | 5.9E-05 | 6.4E-02 | -3.4E-01 | 7.4E-02 | 6.4E-06 |
| rs76326501 | 2 | 43167878 | C | A | 0.0891 | 1.3E-04 | 1.4E-01 | -5.5E-01 | 8.3E-02 | 3.1E-11 |
| rs76452347 | 9 | 35906471 | T | C | 0.2058 | 2.6E-05 | 2.8E-02 | -1.7E-01 | 6.2E-02 | 5.8E-03 |
| rs76785029 | 12 | 94882905 | T | C | 0.0717 | 2.2E-06 | 2.5E-03 | -7.9E-02 | 9.7E-02 | 4.2E-01 |
| rs7777128 | 7 | 27337113 | C | G | 0.0816 | 1.2E-04 | 1.3E-01 | 5.5E-01 | 8.4E-02 | 4.7E-11 |
| rs7810028 | 7 | 139461616 | G | C | 0.1945 | 4.1E-05 | 4.5E-02 | -2.2E-01 | 5.8E-02 | 1.5E-04 |
| rs78378222 | 17 | 7571752 | G | T | 0.0164 | 2.9E-06 | 3.2E-03 | -1.8E-01 | 2.1E-01 | 3.8E-01 |
| rs78648104 | 6 | 50683009 | C | T | 0.1015 | 6.2E-05 | 6.8E-02 | 3.6E-01 | 8.3E-02 | 1.7E-05 |
| rs79089478 | 17 | 40317241 | C | T | 0.0278 | 2.8E-06 | 3.1E-03 | -1.4E-01 | 1.5E-01 | 3.4E-01 |
| rs7914287 | 10 | 69350563 | C | T | 0.2303 | 2.1E-05 | 2.3E-02 | 1.5E-01 | 5.6E-02 | 7.8E-03 |
| rs79146658 | 2 | 179786068 | C | T | 0.0821 | 8.2E-09 | 8.9E-06 | 4.5E-03 | 8.6E-02 | 9.6E-01 |
| rs7927515 | 11 | 76125330 | A | C | 0.3455 | 3.5E-05 | 3.9E-02 | 1.7E-01 | 4.9E-02 | 4.8E-04 |
| rs7977389 | 12 | 49981722 | C | T | 0.1065 | 1.4E-05 | 1.5E-02 | -1.6E-01 | 7.5E-02 | 2.8E-02 |
| rs8059962 | 16 | 81574197 | C | T | 0.5764 | 3.7E-05 | 4.1E-02 | 1.7E-01 | 4.7E-02 | 3.0E-04 |
| rs8105753 | 19 | 31927547 | C | A | 0.3745 | 4.5E-05 | 4.9E-02 | -1.9E-01 | 4.9E-02 | 9.9E-05 |
| rs8258 | 11 | 117283676 | C | T | 0.6324 | 3.9E-06 | 4.3E-03 | -5.6E-02 | 4.8E-02 | 2.4E-01 |
| rs869396 | 4 | 169688000 | A | C | 0.4668 | 4.3E-05 | 4.7E-02 | -1.8E-01 | 4.7E-02 | 1.1E-04 |
| rs871606 | 4 | 54799245 | C | T | 0.1058 | 6.6E-05 | 7.2E-02 | -3.6E-01 | 7.6E-02 | 2.2E-06 |
| rs880315 | 1 | 10796866 | C | T | 0.348 | 3.3E-04 | 3.6E-01 | 5.2E-01 | 5.0E-02 | 1.3E-25 |
| rs8904 | 14 | 35871217 | A | G | 0.3741 | 1.3E-04 | 1.4E-01 | 3.2E-01 | 4.8E-02 | 2.0E-11 |

| SNP | Chromosome number | Chromosome position | Effect allele | Other allele | Effect allele frequency | R-squared | F-statistic | $\beta_{xj}$ | $\beta_{xj}$ Standard error | $\beta_{xj}$ P-value |
|---|---|---|---|---|---|---|---|---|---|---|
| rs891511 | 7 | 150704843 | A | G | 0.3518 | 7.8E-05 | 8.5E-02 | -2.5E-01 | 5.2E-02 | 1.2E-06 |
| rs894344 | 8 | 135612745 | G | A | 0.4105 | 2.8E-05 | 3.0E-02 | 1.5E-01 | 4.7E-02 | 1.8E-03 |
| rs900145 | 11 | 13293905 | T | C | 0.7041 | 1.4E-05 | 1.6E-02 | 1.1E-01 | 5.0E-02 | 2.3E-02 |
| rs917275 | 7 | 28658522 | G | A | 0.3892 | 1.6E-05 | 1.8E-02 | 1.1E-01 | 4.8E-02 | 1.8E-02 |
| rs918466 | 3 | 64710253 | A | G | 0.4105 | 2.5E-05 | 2.7E-02 | -1.4E-01 | 4.8E-02 | 3.6E-03 |
| rs9306160 | 21 | 45107562 | C | T | 0.6026 | 5.5E-05 | 6.0E-02 | 2.1E-01 | 4.7E-02 | 1.3E-05 |
| rs9323988 | 14 | 98587630 | C | T | 0.3896 | 7.2E-05 | 7.9E-02 | 2.4E-01 | 4.6E-02 | 3.1E-07 |
| rs932764 | 10 | 95895940 | G | A | 0.4439 | 1.8E-04 | 1.9E-01 | 3.7E-01 | 4.7E-02 | 4.8E-15 |
| rs9337951 | 10 | 30317073 | A | G | 0.337 | 1.0E-05 | 1.1E-02 | 9.3E-02 | 5.3E-02 | 7.8E-02 |
| rs9349379 | 6 | 12903957 | G | A | 0.4088 | 8.9E-05 | 9.8E-02 | -2.6E-01 | 4.9E-02 | 6.4E-08 |
| rs9372498 | 6 | 118572486 | A | T | 0.0848 | 5.4E-05 | 5.9E-02 | 3.6E-01 | 8.3E-02 | 1.4E-05 |
| rs9479200 | 6 | 152398505 | G | A | 0.1231 | 5.6E-06 | 6.1E-03 | 9.9E-02 | 7.1E-02 | 1.7E-01 |
| rs9549328 | 13 | 113636156 | T | C | 0.2345 | 4.6E-05 | 5.0E-02 | 2.2E-01 | 5.5E-02 | 8.6E-05 |
| rs956006 | 15 | 62808539 | T | C | 0.336 | 5.8E-06 | 6.4E-03 | -7.0E-02 | 4.9E-02 | 1.6E-01 |
| rs9662255 | 1 | 9441949 | A | C | 0.4307 | 4.7E-05 | 5.1E-02 | -1.9E-01 | 4.8E-02 | 9.4E-05 |
| rs9678851 | 2 | 27887034 | A | C | 0.559 | 1.7E-05 | 1.9E-02 | -1.1E-01 | 4.7E-02 | 1.7E-02 |
| rs9687065 | 5 | 148391140 | G | A | 0.1957 | 5.3E-05 | 5.8E-02 | -2.5E-01 | 5.9E-02 | 2.3E-05 |
| rs9729719 | 1 | 38298207 | A | G | 0.2946 | 1.1E-05 | 1.3E-02 | 1.0E-01 | 5.3E-02 | 5.6E-02 |
| rs9810888 | 3 | 53635595 | G | T | 0.5024 | 3.0E-05 | 3.3E-02 | 1.5E-01 | 4.6E-02 | 1.2E-03 |
| rs9815354 | 3 | 41912651 | A | G | 0.1711 | 3.0E-06 | 3.3E-03 | -6.3E-02 | 6.3E-02 | 3.2E-01 |
| rs9827472 | 3 | 56726646 | T | C | 0.3577 | 3.3E-05 | 3.6E-02 | -1.6E-01 | 4.9E-02 | 7.2E-04 |
| rs9888615 | 14 | 53377540 | C | T | 0.7064 | 6.2E-05 | 6.7E-02 | 2.4E-01 | 5.0E-02 | 2.3E-06 |

**eTable 3. UK prevalence estimates used to calculate population impact of distributional shifts in BP**

| Outcome | UK Prevalence (per 10,000) | Age group (years) | Year | Source of prevalence estimate |
|---|---|---|---|---|
| **Coronary Artery Disease** | 612 | 45-64 | 2017 | Health Survey for England [1] |
| **Ischemic Cerebrovascular Disease (All)** | 280 | 45-64 | 2017 | Health Survey for England [1] |
| **Hemorrhagic Stroke (All)** | 84.7 | 50-69 | 2010 | Global Burden of Disease [2] |
| **Peripheral Vascular Disease** | 240 | 50-89 | 2014 | The Health Improvement Network [3] |
| **Aortic Valve Stenosis** | 96.0 | 45-64 | 2017 | Health Survey for England [1] |
| **Atrial Fibrillation** | 250 | 45-69 | 2017 | Public Health England [4] |
| **Heart failure** | 80.0 | 45-74 | 2017 | Quality and Outcomes Framework [5] |
| **Dilated Cardiomyopathy** | 20.0 | 45-75 | 2014 | British Heart Foundation [6] |
| **Endocarditis** | 11 | All ages[a] | 2013 | Hospital Episode Statistics [7] |
| **Rheumatic Heart Disease** | 23.0 | All ages[a] | 2013 | Hospital Episode Statistics [8] |
| **Chronic Kidney Disease** | 700 | 45-64 | 2017 | Health Survey for England [1] |

[a] When no age group-specific estimates could be obtained, the total population prevalence was used

**eTable 4. Mendelian randomization estimates (odds ratio with 95% confidence interval per 10 mmHg increase in genetically-predicted SBP) using the inverse-variance weighted, MR-Egger, weighted median, and MR-PRESSO methods for 21 disease outcomes.**

| Outcome | Inverse-weighted variance | | MR-Egger | | Weighted median | | MR-PRESSO | |
|---|---|---|---|---|---|---|---|---|
| | OR (95% CI) | P-value | OR (95% CI) | P-value | OR (95% CI) | P-value | OR (95% CI) | P-value |
| **Coronary Artery Disease** | 1.59 (1.45-1.74) | <0.001 | 1.44 (1.20-1.73) | <0.001 | 1.61 (1.48-1.75) | <0.001 | 1.59 (1.45-1.75) | <0.001 |
| **Ischemic Cerebrovascular Disease (all)** | 1.52 (1.39-1.66) | <0.001 | 1.66 (1.40-1.98) | <0.001 | 1.50 (1.33-1.70) | <0.001 | 1.52 (1.39-1.66) | <0.001 |
| **Ischemic Stroke** | 1.70 (1.52-1.90) | <0.001 | 1.86 (1.48-2.32) | <0.001 | 1.78 (1.51-2.09) | <0.001 | 1.71 (1.52-1.91) | <0.001 |
| **Transient Ischemic Attack** | 1.34 (1.19-1.51) | <0.001 | 1.40 (1.10-1.79) | 0.006 | 1.38 (1.15-1.64) | <0.001 | 1.35 (1.19-1.52) | <0.001 |
| **Hemorrhagic Stroke (all)** | 1.43 (1.21-1.69) | <0.001 | 1.39 (0.99-1.95) | 0.05 | 1.33 (1.04-1.71) | 0.02 | 1.42 (1.20-1.68) | <0.001 |
| **Intracerebral Hemorrhage** | 1.32 (1.06-1.66) | 0.02 | 1.22 (0.78-1.91) | 0.39 | 1.24 (0.88-1.74) | 0.21 | 1.31 (1.04-1.64) | 0.02 |
| **Subarachnoid Hemorrhage** | 1.54 (1.22-1.94) | <0.001 | 1.72 (1.09-2.72) | 0.02 | 1.65 (1.17-2.32) | 0.004 | 1.53 (1.22-1.94) | <0.001 |
| **Aortic Aneurysm (all)** | 1.26 (1.02-1.56) | 0.03 | 1.51 (0.99-2.31) | 0.06 | 1.39 (1.06-1.82) | 0.02 | 1.28 (1.03-1.58) | 0.02 |
| **Abdominal Aortic Aneurysm** | 1.20 (0.93-1.56) | 0.17 | 1.07 (0.64-1.79) | 0.80 | 1.34 (0.95-1.88) | 0.10 | 1.22 (0.94-1.58) | 0.15 |
| **Thoracic Aortic Aneurysm** | 1.19 (0.79-1.77) | 0.41 | 1.99 (0.90-4.41) | 0.09 | 1.34 (0.75-2.40) | 0.33 | 1.23 (0.82-1.84) | 0.31 |
| **Venous Thromboembolism (all)** | 0.90 (0.80-1.01) | 0.07 | 1.00 (0.79-1.25) | 0.98 | 0.87 (0.78-0.96) | 0.005 | 0.90 (0.80-1.01) | 0.07 |
| **Deep Vein Thrombosis** | 0.88 (0.77-1.00) | 0.04 | 0.95 (0.74-1.22) | 0.67 | 0.90 (0.80-1.02) | 0.09 | 0.87 (0.77-0.99) | 0.04 |
| **Pulmonary Embolism** | 0.91 (0.79-1.05) | 0.20 | 1.05 (0.79-1.38) | 0.75 | 0.96 (0.83-1.11) | 0.57 | 0.92 (0.80-1.05) | 0.22 |
| **Peripheral Vascular Disease** | 1.28 (1.11-1.46) | <0.001 | 1.01 (0.77-1.32) | 0.96 | 1.18 (0.98-1.43) | 0.09 | 1.28 (1.11-1.47) | <0.001 |
| **Aortic Valve Stenosis** | 1.74 (1.48-2.04) | <0.001 | 1.90 (1.38-2.62) | <0.001 | 1.81 (1.43-2.29) | <0.001 | 1.75 (1.49-2.05) | <0.001 |
| **Atrial Fibrillation** | 1.32 (1.21-1.42) | <0.001 | 1.35 (1.15-1.58) | <0.001 | 1.29 (1.17-1.42) | <0.001 | 1.32 (1.21-1.43) | <0.001 |
| **Heart Failure** | 1.38 (1.25-1.53) | <0.001 | 1.36 (1.12-1.67) | 0.002 | 1.47 (1.28-1.69) | <0.001 | 1.37 (1.24-1.51) | <0.001 |
| **Dilated Cardiomyopathy** | 1.61 (1.24-2.10) | <0.001 | 2.57 (1.52-4.35) | <0.001 | 1.60 (1.07-2.40) | 0.02 | 1.58 (1.23-2.03) | <0.001 |
| **Endocarditis** | 1.49 (1.12-1.99) | 0.007 | 1.41 (0.79-2.52) | 0.24 | 1.47 (0.97-2.24) | 0.07 | 1.52 (1.14-2.03) | <0.001 |
| **Rheumatic Heart Disease** | 1.32 (1.13-1.53) | <0.001 | 1.55 (1.15-2.09) | 0.004 | 1.43 (1.16-1.77) | 0.001 | 1.31 (1.13-1.52) | <0.001 |
| **Chronic Kidney Disease** | 1.39 (1.24-1.55) | <0.001 | 1.17 (0.95-1.46) | 0.14 | 1.38 (1.19-1.59) | <0.001 | 1.39 (1.24-1.55) | <0.001 |

**eTable 5. Population impact fractions (PIFs) for outcomes with strong evidence of causality for SBP. The PIF represents the percentage reduction (with 95% confidence interval) in events if SBP was 132.7 mmHg, 127.7 mmHg, and 115.0 mmHg for all individuals in the UKBB study sample, instead of the current mean SBP of 137.7 mmHg.**

| Outcomes | Percentage reduction in events attributable to setting SBP to: | | |
|---|---|---|---|
| | 132.7 mmHg (-5 mmHg) | 127.7 mmHg (-10 mmHg) | 115.0 mmHg (-22.7 mmHg) |
| **Coronary Artery Disease** | 20.9 (17.3-24.4) | 37.5 (31.6-42.9) | 65.6 (57.8-72.0) |
| **Ischemic Cerebrovascular Disease (all)[a]** | 18.9 (15.2-22.5) | 34.3 (28.1-40.0) | 61.5 (52.7-68.6) |
| **Hemorrhagic Stroke (all)[a]** | 16.1 (8.6-22.9) | 29.5 (16.5-40.5) | 54.8 (33.5-69.3) |
| **Peripheral Vascular Disease** | 11.8 (5.4-17.7) | 22.1 (10.4-32.3) | 43.3 (22.1-58.7) |
| **Aortic Valve Stenosis** | 24.4 (18.1-30.2) | 42.9 (33.0-51.3) | 72.0 (59.7-80.5) |
| **Atrial Fibrillation** | 12.6 (9.1-16.5) | 23.7 (17.3-30.2) | 45.8 (35.0-55.8) |
| **Heart Failure** | 14.4 (10.0-18.5) | 26.7 (18.9-33.6) | 50.5 (37.9-60.6) |
| **Dilated Cardiomyopathy** | 20.5 (9.1-30.2) | 36.9 (17.3-51.3) | 64.8 (35.0-80.5) |
| **Endocarditis** | 18.9 (6.3-29.9) | 34.2 (12.2-50.8) | 61.4 (25.6-80.0) |
| **Rheumatic Heart Disease** | 12.6 (5.8-18.9) | 23.7 (11.3-34.3) | 45.8 (23.9-61.5) |
| **Chronic Kidney Disease** | 15.2 (10.4-19.8) | 28.1 (19.8-35.7) | 52.7 (39.3-63.2) |
| **Total** | **16.9 (12.2-21.3)** | **30.8 (22.8-38.0)** | **56.2 (43.7-65.9)** |

[a] To minimize double counting, we present single estimates for both ischemic cerebrovascular disease and hemorrhagic stroke
